# Large-scale Integrative Analysis of Juvenile Idiopathic Arthritis for New Insight into Its Pathogenesis

**DOI:** 10.1101/2023.04.07.23287912

**Authors:** Daeun Kim, Jaeseung Song, Nicholas Mancuso, Serghei Mangul, Chul Woo Ahn, Junghyun Jung, Wonhee Jang

## Abstract

**Objectives:** Juvenile idiopathic arthritis (JIA) is one of the most prevalent rheumatic disorders in children and is classified as an autoimmune disease (AID). While a robust genetic contribution to JIA etiology has been established, the exact pathogenesis remains unclear. We conducted a comprehensive integrative analysis to gain new insights into the etiology of JIA.

**Methods:** To prioritize biologically interpretable susceptibility genes and proteins for JIA, we conducted transcriptome-wide and proteome-wide association studies (TWAS/PWAS). Then, to understand genetic architecture JIA, we systematically analyzed single nucleotide polymorphism (SNP)-based heritability, a signature of natural selection, and polygenicity. Finally, we performed HLA typing using multi-ancestry RNA sequencing data and analyzed the T cell receptor (TCR) repertoire at a single-cell level to investigate the associations between immunity and JIA risk.

**Results:** We have identified 19 TWAS genes and two PWAS proteins that are associated with JIA risks. Furthermore, we observe that the heritability and cell type enrichment analysis of JIA are enriched in T lymphocytes and HLA regions, and that JIA shows higher polygenicity compared to other AIDs. In multi-ancestry HLA typing, B*45:01 is more prevalent in African JIA patients than in European JIA patients, whereas DQA1*01:01, DQA1*03:01, and DRB1*04:01 exhibit a higher frequency in European JIA patients. Using single-cell immune repertoire analysis, we identify clonally expanded T cell subpopulations in JIA patients, including *CXCL13*^+^*BHLHE40*^+^ T_H_ cells which are significantly associated with JIA risks.

**Conclusions:** Our findings shed new light on the pathogenesis of JIA and provide a strong foundation for future mechanistic studies aimed at uncovering the molecular drivers of JIA

## Introduction

Juvenile idiopathic arthritis (JIA) is one of the most common rheumatic diseases in children; it is mainly regarded as an autoimmune disease (AID) whose clinical manifestations include persistent limping, painful joints, stiffness, and inflammation^1, 2^. The exact JIA pathogenesis remains unclear; however, it has been demonstrated that there is a strong genetic contribution to the etiology of JIA. The single nucleotide polymorphism (SNP)-based heritability for JIA was estimated to be 73%, among the most highly heritable pediatric AIDs. Even though genome-wide association studies (GWASs) have identified many risk variants of JIA, most are located in non-coding regions, making it difficult to interpret their functional consequences^3^. By integrating GWASs with expression quantitative trait loci (eQTL), transcriptome-wide association studies (TWASs) provide a powerful approach to prioritize susceptibility genes affected by risk variants^4^.

Investigating the genetic architecture of complex diseases is essential for understanding the genetic basis of phenotypic variations and evolutions^5^. For complex diseases, natural selection plays an essential role in forming the genetic architecture and provides valuable insights into biological mechanisms^6^. In AIDs, the signature of negative selection is significant because it sheds light on how the human immune system has evolved to defend against pathogens while avoiding harmful responses against self-tissues.

Moreover, given that JIA is considered an AID, it is also important to further understand the underlying mechanisms of immune responses affecting the JIA etiologies. Human leukocyte antigen (HLA) molecules presenting peptide antigens to receptors on T lymphocytes are highly polymorphic at their peptide-binding site and mediate the adaptive immune responses^5^. Moreover, T cell receptors (TCRs) interacting with HLA molecules are essential components of adaptive immune responses^7^. Through the somatic recombination of TCRs, self-reactive T cells can be produced, and the recognition of self-antigens can affect the development of AIDs^8^. Even though there is some evidence that T cells are involved in JIA etiologies, their contributions to the pathogenesis of AIDs including JIA have not been completely revealed^9, 10^.

Herein, we prioritized susceptibility genes/proteins for JIA by conducting a multi-tissue TWAS and proteome-wide association study (PWAS) by integrating JIA GWAS summary statistics data (*n* = 3,305 cases and *n* = 9,196 controls) with reference eQTL/protein QTL (pQTL) panels (*n* = 14,037). We identified 19 genes and two proteins associated with JIA risk using TWAS and PWAS. We then estimated disease heritability and signatures of natural selection to understand the genetic architecture and to prioritize the most relevant tissue and cell types of JIA. We observed that the heritability of JIA was enriched in T lymphocytes and HLA regions and that JIA showed higher polygenicity than other AIDs. Next, HLA typing was conducted using multi-ancestry RNA sequencing (RNA-seq) data, and TCR repertoire analysis was performed at a single-cell level to investigate the associations between immunity and JIA risks. We found some HLA types, such as B*45:01, DQA1*01:01, DQA1*03:01, and DRB1*04:01, which were more frequent in the European or African JIA patients. In addition, we identified clonally expanded T cell subpopulations in JIA patients, among which *CXCL13*^+^*BHLHE40*^+^ T cells were significantly associated with JIA risks at the single-cell level. We believe that these findings could provide new insights into understanding the underlying mechanisms of JIA etiology.

## Methods

### Genome-wide association summary statistics of JIA

GWAS summary statistics data of JIA were retrieved from the GWAS catalog and the most recent dataset was used for this study (catalog ID: GCST90010715)^11, 12^. Details on the process of genotyping and quality control were described by López-Isac et al.^12^. The summary statistics of JIA were computed only for the European population (*n* = 3,305 patients and *n* = 9,196 control subjects) and resulted in 6,334,221 SNPs with minor allele frequency (MAF) ≥ 1%. The JIA summary statistics file was converted into a sumstats-formatted file by LD score (LDSC) software (v1.0.1)^13^.

### Transcriptome-wide association study and proteome-wide association study

To identify susceptibility genes associated with the pathogenesis of JIA, a TWAS was performed using functional summary-based imputation (FUSION) (see URLs)^4^. Briefly, TWAS identifies risk genes associated with the target disease by integrating GWAS summary statistics data with the reference eQTL data of specific tissues, considering linkage disequilibrium (LD) structures. Using the eQTL panels and LD information, the *cis*-genetic components of gene expression are imputed from the JIA summary statistics data. Then, the predicted gene expression is used for association tests with JIA risks to identify significant associations between the gene expression and the disease. Twelve connective tissue panels from the Genotype-Tissue Expression project v7 (GTEx v7; *n* = 449), the Metabolic Syndrome in Men study (METSIM; *n* = 563), the Netherlands Twin Registry (NTR; *n* = 1,247), and the Young Finns Study (YFS; *n* = 1,264) were selected as expression weights for transcriptomic imputation (Table S1)^14–19^. For each panel, predictive models for gene expression were trained using *cis*-regulated genes by SNPs within +/−500kb of the transcription start site and are significant for heritability (*cis*-*h*^2^) with P < 0.01. The European LD information from the 1000 Genomes project accounted for the LD regions^20^. Due to the complex LD patterns, we excluded the TWAS associations from major histocompatibility complex (MHC) regions^21^. The significance threshold of TWAS associations was corrected with the Bonferroni correction (P_TWAS_ < 7.55 × 10^−07^, 0.05/66,196).

We conducted a PWAS using the pQTL data from the INTERVAL (*n* = 3,301)^22^ and Atherosclerosis Risk in Communities (ARIC; *n* = 7,213) study^23^ of European individuals. For each pQTL panel, predictive models were trained using *cis*-regulatory proteins with SNPs within +/− 500 Kb of the same transcriptional start site, and the significance levels of *cis*-*h*2 in INTERVAL (*n* = 1,031 models) and ARIC (*n* = 1,309 models) were 0.05 and 0.01, respectively. The predictive models in the INTERVAL study were fitted using the sum of single effects (SuSiE)^24^, and the elastic net was used in the ARIC study. As with TWAS, the PWAS associations in the MHC region were excluded. The same significance thresholds with the Bonferroni correction were used for the PWAS associations as the TWAS associations (P_PWAS_ < 2.16 × 10^−05^, 0.05/2,311).

### Fine-mapping of TWAS and PWAS associations

We performed fine-mapping of causal gene sets (FOCUS) (see URLs) to identify TWAS/PWAS associations responsible for the disease, estimating gene-trait associations at the GWAS risk regions while considering LD structures and controlling for pleiotropic SNP effects^25^. FOCUS calculates the posterior inclusion probability (PIP) per the TWAS/PWAS association and suggests credible gene sets containing susceptibility genes at a 90%-confidence level. We applied FOCUS to the specific loci for each panel where the significant TWAS/PWAS associations (P_TWAS_ < 7.55 × 10^−07^ and P_PWAS_ < 2.16 × 10^−05^) identified by FUSION were detected. The weight database for FOCUS was generated from the GTEx and INTERVAL FUSION weights.

### Pathway enrichment analysis

We conducted a TWAS-based Gene Set Enrichment Analysis (TWAS-GSEA) (see URLs) using the TWAS results from individual tissue panels^26^. The TWAS associations with a panel and the 12 eQTL panels containing information on the position of genes were used as inputs for this analysis. We retrieved curated gene sets from canonical pathways (CP), WikiPathways, Kyoto Encyclopedia of Genes and Genomes (KEGG), BioCarta, Reactome, and Pathway Interaction Database (PID) from the Molecular Signatures Database (MSigDB v7.2) (see URLs)^27–32^.

A GWAS-based pathway enrichment analysis was carried out using the multi-marker analysis of genomic annotation (MAGMA v1.07) (see URLs)^33^. The SNPs were annotated to the corresponding genes, based on dbSNP v151 SNP locations for the European group and NCBI Build 37 gene definitions^20, 34^. Gene sets retrieved from the CPs of MSigDB were used^27–32^.

### Genetic correlation analyses with other traits

To estimate genetic correlations at the genome-wide level between JIA and other AID-like (*n* = 10) and non-AID-like (*n* = 15) traits, an LD score regression was performed using the GWAS summary statistics data of JIA and 25 traits using LDSC^13, 35^. Sumstats-formatted publicly available summary statistics (PASS) data of 22 traits, excluding type 1 diabetes (T1D), were retrieved from LD Hub (see URLs)^36^. The summary statistics data of T1D were obtained from the GWAS catalog (catalog ID: GCST005536) because T1D PASS data were not reported in the LD hub, although the comorbidity of JIA and T1D has previously been reported^11, 37, 38^. Summary statistics data of AID_ALL_ and AID_SURE_ traits, the remaining two traits, reported in UK Biobank (UKBB) (see URLs) were also used. The LD score data for the European samples were used for this analysis^20^.

To estimate transcriptome-wide genetic correlations between JIA and the 23 traits, the TWAS results of these traits were retrieved from the TWAS-hub (see URLs)^4^. Eighteen of the 23 traits were obtained from the TWAS-hub and TWAS analyses of T1D, AID_ALL_, and AID_SURE_ traits were conducted using the same procedure as JIA. Transcriptome-wide genetic correlations between JIA, T1D, AID_ALL_, AID_SURE_, and the 18 traits were estimated using the RHOGE (see URLs)^39^.

### Weighted gene co-expression network analysis

This study used an Affymetrix microarray dataset (GEO study: GSE13501^40^) and two RNA-seq datasets (GEO study: GSE112057^41^ and GSE79970^42^) containing mRNA expression data on JIA patients and healthy controls. The preprocessing and normalization of datasets were performed in accordance with Kim et al.^2^. The top 7,000 most-expressed genes of the GSE13501^40^ dataset were selected for simplicity after normalization following Jung et al.^43^. A signed weighted gene co-expression network analysis (WGCNA) (see URLs) was performed to identify co-expression modules comprising positively correlated genes based on bi-weight mid-correlation^44^. A soft-thresholding power (β) of seven was selected for the network construction (scale-free *r*^2^ = 0.8). The expression profile of each module was summarized by the module eigengene (ME). The minimum size of modules was 50 genes and paired modules with high ME (r > 85) were merged. With the co-expression modules, module preservation analyses were conducted using the microarray dataset as a reference set and each RNA-seq dataset as a test set with the co-expression modules^45^. The analyses were permuted up to 1,000 times and Z-summary scores were computed to identify the preserved modules. To examine whether TWAS associations were enriched in the co-expression modules, GSEA was performed with the fgsea R package (see URLs) using the co-expression modules as the reference gene sets^46^. The TWAS associations from each panel, arranged by their Z-scores in descending order, were used as the pre-ranked gene sets. Functional annotation of the co-expression modules, in which TWAS associations were enriched, was carried out using the Database for Annotation, Visualization, and Integrated Discovery (DAVID) (see URLs)^47^.

### JIA-relevant-tissue and cell-type analyses

LD score regression to specifically expressed genes (LDSC-SEG) v1.0.1 (see URLs) was applied to determine disease-relevant tissues and cell types in JIA^48^. Two types of precomputed expression datasets were downloaded for the analysis: 1) human RNA-seq data of 53 tissues/cell types from the GTEx^49^ and human/mouse/rat array data of 152 tissues/cell types from Franke lab^50, 51^ and 2) mouse array data of 292 immune cell types from ImmGen^52^. Two types of precomputed chromatin datasets were also downloaded: 1) human 431 tissue-specific epigenomic annotations from peaks for six epigenetic marks from Roadmap Epigenomics^53^ and ENCODE projects^54^ and 2) human ATAC-seq peaks from 13 cell types for human hematopoietic hierarchy^55^. Fetal data was excluded from the epigenomic data. The Bonferroni correction was applied to determine the significance levels.

### Estimating the SNP-based heritability and expression-mediated heritability

We estimated SNP-based SNP heritability (□^2^_□□□_) in a set of 1,702 independently partitioned genomic blocks^56^ across the genome using Heritability Estimation from Summary Statistics (HESS)^39^ v0.5.3-beta (see URLs). To account for the large number of hypotheses tested, the Bonferroni correction was performed at α = 0.05/1702 to determine significant levels. The mediated expression score regression (MESC)^57^ was used to estimate the proportion of heritability mediated by a *cis*-genetic component of gene expression levels (□_□_). The MESC software was downloaded, along with its precomputed expression scores from the GTEx consortium and eQTLGen^58^.

### Genetic architecture analysis

To identify signatures of negative selection for JIA, we utilized summary-data-based BayesS (SBayesS)^59^ in the GCTB software (see URLs) by estimating the joint posterior distribution of effect size and MAF. Based on the Markov-chain Monte Carlo (MCMC) sample, the posterior mean was used as a point estimation, and the posterior standard error was approximated by the standard deviation. Moreover, a sparse LD matrix was used for computational efficiency^59^.

### Estimation of HLA gene expression and HLA typing analysis

Consensus HLA typing analysis was carried out using the GSE112057^41^ RNA-seq dataset by seven HLA-typing software (see URLs): seq2HLA (v2.3), arcasHLA (v0.2.0), HLAforest, HLA-VBSeq (v2), OptiType (v1.3.3), PHLAT (v1.0), and HLA typing from the high-quality dictionary (HLA-HD) (v1.2.1)^42, 60–66^. The GSE112057^41^ dataset contains expression data on 115 JIA patients (43 oligo-JIA, 46 poly-JIA, and 26 sJIA; 34 African JIA and 81 European JIA) and 12 control subjects. The reads mapped to chromosome 6 were detected by STAR software^67^ (v2.5.3a) (see URLs) using the human reference genome (hg19) and were then analyzed by seven HLA-typing software with default settings. The International Immunogenetics Project/HLA database^68, 69^ (v3.10.0) was used for arcasHLA, HLA-VBSeq, and HLA-HD. The consensus frequencies of HLA allele types (HLA class I: A, B, and C; HLA class II: DP alpha 1 [DPA1], DP beta 1 [DPB1], DQ alpha 1 [DQA1], DQ beta 1 [DQB1], DR alpha [DRA], DR beta 1 [DRB1], DRB3, DRB4, and DRB5) were calculated using the HLA typing result per the software at two-field resolution as a replica and integrating the seven replicates, grouped by disease states (control and JIA). The same procedure was also performed using the GSE112057^41^ grouped by disease subtypes (healthy control, oligo-JIA, poly-JIA, and sJIA) and ancestry (healthy control, African JIA, and European JIA).

The expression levels of HLA genes were estimated by seq2HLA^60^ using the GSE112057^41^ dataset. FASTQ-formatted files in GSE112057^41^ were used as input files and the locus-specific expression levels of HLA genes were measured as read per kilobase million (RPKM). HLA class I and II gene expression levels were compared between JIA and control groups.

### Profiling adaptive immune repertoires in JIA

The unmapped reads were extracted from the GSE112057^41^ dataset following a read origin protocol (ROP)^70^ (see URLs). SRR6868722 and SRR6868696 were excluded due to quality problems. To analyze the TCR repertoires, the reads mapped onto the complementary determining region 3 (CDR3) in TCR loci were identified using immune profiling by ROP (ImReP)^71^ (see URLs). ImReP assembles the clonotypes, defined as clones having identical CDR3 amino-acid sequences, and identifies the corresponding V(D)J recombination. The alpha diversity (Shannon entropy) of TCRs was measured within the immune repertoire of an individual. The alpha diversities of TCRs (TCR α and β) were respectively compared between control and JIA groups. The same analyses were conducted using the GSE112957^41^ dataset grouped by JIA subtypes (healthy controls, oligo-JIA, poly-JIA, and sJIA).

### Single-cell analysis

The single-cell RNA-seq (scRNA-seq) of JIA were downloaded from NCBI SRA (SRA study: SRP288574^72^). The 10x Genomics BAM file contains single-cell transcriptome and TCR repertoire data from single CD4^+^CD45RO^+^CD25^−^ (CD4^+^) and CD8^+^CD45RO^+^ (CD8^+^) T cells of synovial fluid (SF) and peripheral blood (PB) tissue in seven oligo-JIA patients. The BAM files mapped to human hg19 references were converted into FASTQ files using the bamtofastq (v1.3.1) tool and remapped to human GRCh38 references (ref-2020-A). The FASTQ data were processed using the cellranger^73^ (v6.1.2) count tools and analyzed using the Seurat^74^ R package. To preprocess the data, we filtered out cells with either >4,000 or <200 distinct features and those with >10% mitochondrial count. We conducted normalization, identification of highly variable features (i.e., feature selection), and scaling data with default settings. After performing dimensional reduction using Principal Component Analysis (PCA), we used the Harmony^75^ R package to integrate datasets derived from seven patients and two tissue types. Based on the 40 components of Harmony, we carried out Uniform Manifold Approximation and Projection (UMAP)^76^ and nearest-neighbor graph construction. Then, we determined single-cell clusters using 0.5 resolution. We retrieved the results of the single-cell TCR repertoire profiling from NCBI GEO (GEO study: GSE160097).

### Statistical analysis

The Bonferroni correction was applied to determine the significance thresholds of TWAS associations (P_TWAS_ < 7.55 × 10^−07^, 0.05/66,196) and PWAS associations (P_PWAS_ < 2.16 × 10^−05^, 0.05/2,311). The significance thresholds with the Bonferroni correction were also used in the LDSC-SEG analysis, heritability enrichment analysis, and genetic architecture analysis. A false discovery rate (FDR) was used to determine the significance threshold for the TWAS-GSEA, the MAGMA gene set analysis, the genetic correlation analyses, and the functional annotation analysis using DAVID. The consensus frequencies of HLA allele types were compared using two-sided 2-(control and JIA), 3-(control, African JIA, and European JIA), and 4-sample (control, oligo-JIA, poly-JIA, and sJIA) proportion tests, respectively. Using a two-tailed t-test, HLA class I and II gene expression levels were compared between control and JIA groups. The alpha diversities of TCRs were compared between control and JIA groups using a two-tailed t-test. When grouped by JIA subtypes (control, oligo-JIA, poly-JIA, and sJIA), the alpha diversities of TCRs were compared using one-way ANOVA with post hoc Tukey HSD.

## Results

### Identification of susceptibility genes associated with JIA risk using TWAS

To prioritize susceptibility genes and proteins for JIA risk, we performed a TWAS by integrating JIA GWAS data with the predicted expression of 66,196 gene/tissue pairs from 12 eQTL datasets. We focused on eQTL derived from connective tissues due to JIA reflecting a chronic inflammation of connective tissues (Table S2)^4,77^. We identified 35 significant TWAS associations across 19 genes in four independent genomic regions (1p13.2, 1q21.3, 5q11.2, and 16p11.2-12.1) (P_TWAS_ < 7.55 × 10^−07^; Fig. 1 and Table S3). Excluding three non-coding genes and one pseudogene (*NPIPB7*) from the 19 TWAS genes, 11 out of 15 (73%) genes have been suggested as JIA-associated genes, which confirms that our results were consistent with previous studies^3,12, 78–80^. Among the remaining four genes, *MAGI3* and *NFATC2IP* were mentioned by a previous JIA TWAS study^81^, while *DCLRE1B* and *NPIPB9* have not previously been emphasized as susceptibility genes for JIA, to the best of our knowledge (Fig. 1). Next, we conducted a PWAS using predictive models of plasma proteins from INTERVAL^22^ and ARIC^23^. We observed two significant PWAS associations of IL27 and ERAP2 in two genomic regions (16p11.2-12.1 and 5q15) (P_PWAS_ < 2.16 × 10^−05^; Fig. 1 and Table S4), which were reportedly involved in JIA risks^12, 81^.

**Figure 1.**
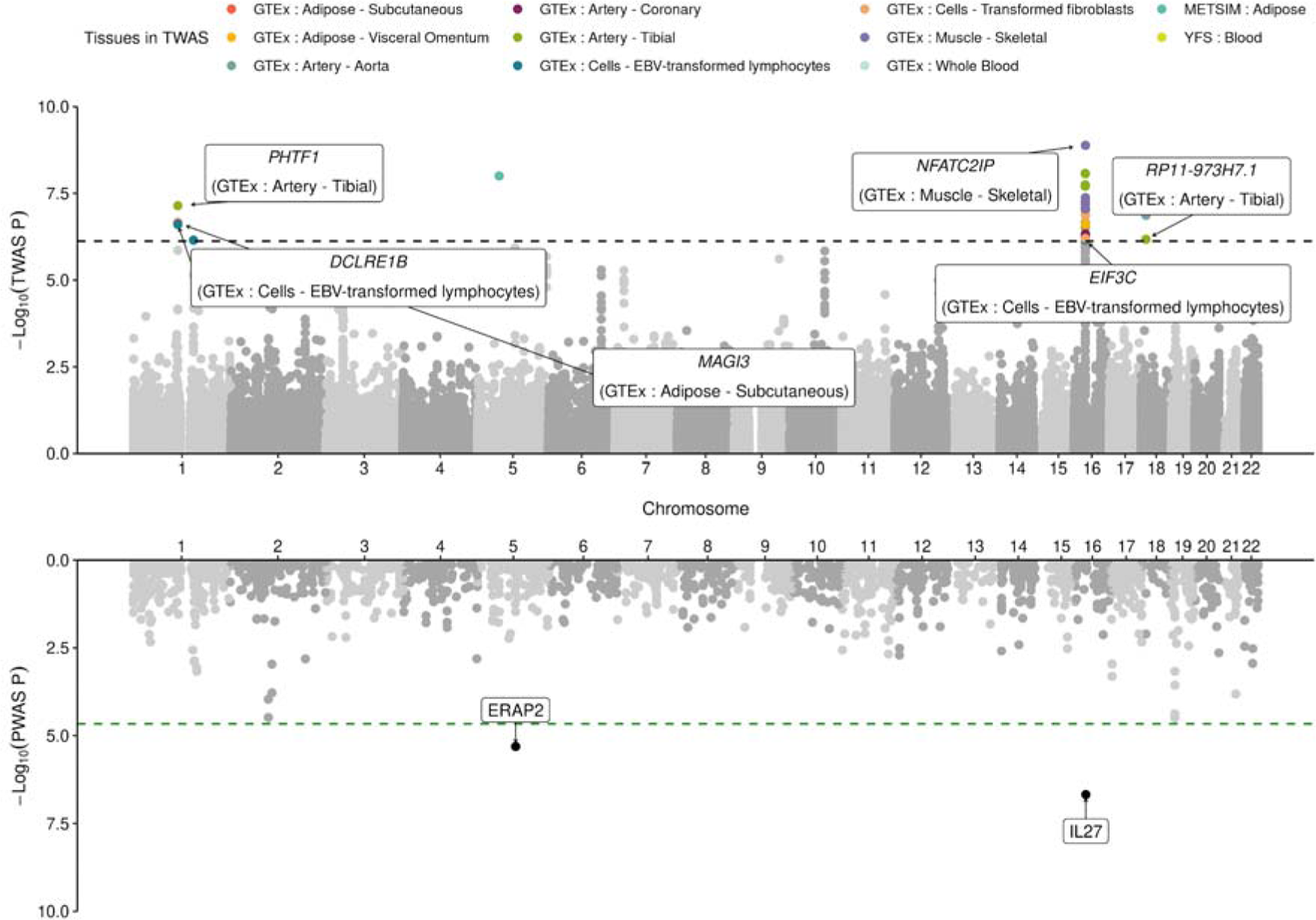
Manhattan plots for JIA TWAS/PWAS result. The upper and lower panels show Manhattan plots for JIA TWAS and PWAS results, respectively. Each dot represents a P-value for a TWAS or PWAS association between JIA and the predicted expression level of a gene or *cis*-regulated plasma protein. The black and green horizontal dashed lines indicate the significance thresholds of TWAS and PWAS with Bonferroni correction (P_TWAS_ < 7.55 × 10^−07^ and P_PWAS_ < 2.16 × 10^−05^). Statistically significant TWAS associations of JIA are colored by tissue panels. The black dots of the lower panel indicate statistically significant PWAS associations of JIA. The names of TWAS genes and PWAS proteins with PIP > 0.2 in FOCUS are labeled.

To distinguish between genes likely causal for JIA risk from those that tag risk due to LD and shared regulatory features, we performed a probabilistic fine-mapping analysis by FOCUS^25^ using the same expression weights used in our FUSION analyses. Among the 19 TWAS genes identified by FUSION, FOCUS identified nine genes in 90%-credible sets, which we denote as putatively causal genes affected by genome-wide significant GWAS signals (Table S5). Six of the nine (67%) genes had >0.4 PIPs, of which *MAGI3* (PIP = 0.5), *NFATC2IP* (PIP = 0.417), and *DCLRE1B* (PIP = 0.405) were putatively responsible for JIA risk. Similarly, fine-mapping of PWAS results prioritized IL27 with a PIP of 0.98 in the plasma protein from the INTERVAL study (Table S5). ERAP2 from the ARIC study was included in credible sets with a PIP of 0.23. Altogether, we identified 19 susceptibility genes/ two risk proteins for JIA using TWAS/PWAS and prioritized likely causal genes for JIA risk, suggesting that our findings provide novel insights into the etiology of JIA.

### Exploring biological pathways that may contribute to the pathogenesis of JIA

To explore the biological effects derived from overall TWAS associations from the multi-tissue panels, we conducted GSEA with the JIA TWAS results using CP gene sets^26^. We found a total of nine CP gene sets were significantly involved with JIA TWAS associations (FDR < 0.05). Three of the nine gene sets are characterized by sulfation (Table 1). The impaired sulfation pathway was reportedly implicated in diastrophic dysplasia leading to cartilage disorder and joint degradations, similar to the clinical manifestations of JIA^82^. In addition, tyrosine-sulfated proteins were reported to play roles in the pathogenesis of various AIDs^83^. Four other gene sets involved in the IL27 pathway, IL6 family signaling, nitric oxide 2 IL12 (NO2IL12) pathway, and IL17 pathway are associated with immune responses that are representatives of AIDs. The T cell apoptosis pathway was also significantly implicated in JIA associated with the dysregulated T cell responses^84^. Moreover, stathmin is known to play a critical role in regulating the cell cycle, and its phosphorylation is reportedly important in T cell activation^85, 86^.

**Table 1.** Biological pathways significantly involved in JIA based on the TWAS associations

| Gene set | Panel | P-value | FDR |
| --- | --- | --- | --- |
| Wikipathway: Sulfation biotransformation reaction | GTEX: Cells - Transformed fibroblasts | $4.56 \times 10^{-07}$ | $8.41 \times 10^{-04}$ |
| | GTEX: Muscle - Skeletal | $7.58 \times 10^{-06}$ | $1.12 \times 10^{-02}$ |
| Reactome: Cytosolic sulfonation of small molecules | GTEX: Cells - Transformed fibroblasts | $6.11 \times 10^{-06}$ | $5.63 \times 10^{-03}$ |
| KEGG: Sulfur metabolism | GTEX: Muscle - Skeletal | $1.16 \times 10^{-05}$ | $1.12 \times 10^{-02}$ |
| PID: IL27 pathway | GTEX: Whole - Blood | $4.97 \times 10^{-07}$ | $8.38 \times 10^{-04}$ |
| Reactome: Interleukin 6 family signaling | GTEX: Whole - Blood | $1.65 \times 10^{-06}$ | $1.39 \times 10^{-03}$ |
| BioCarta: NO2IL12 pathway | GTEX: Whole - Blood | $1.10 \times 10^{-05}$ | $6.17 \times 10^{-03}$ |
| | YFS: Blood | $3.09 \times 10^{-05}$ | $2.33 \times 10^{-02}$ |
| BioCarta: T-cell apoptosis (TCAPOPTOSIS) pathway | YFS: Blood | $9.47 \times 10^{-06}$ | $1.85 \times 10^{-02}$ |
| BioCarta: Stathmin pathway | YFS: Blood | $3.57 \times 10^{-05}$ | $2.33 \times 10^{-02}$ |
| BioCarta: IL17 pathway | YFS: Blood | $5.82 \times 10^{-05}$ | $2.84 \times 10^{-02}$ |

We additionally performed a GWAS-based pathway enrichment analysis using MAGMA^33^ with JIA GWAS summary statistics and CP gene sets, given that HLA signals were dropped from TWAS-based results due to complicated LD. The results showed that a total of 40 gene sets were significantly implicated in JIA (FDR < 0.05; Table S6). Most pathways were associated with various immune system components, such as inflammatory cytokines, HLA, immunoglobulins, and T cells. The IL27 pathway, IL6 family signaling, NO2IL12 pathway, and IL17 pathway were observed in both TWAS-GSEA and MAGMA (Tables 2 and S8). The gene sets representing T1D and autoimmune thyroid disease (AITD) were also related to JIA, supporting the idea that JIA is indeed an AID. Collectively, our results suggest that impaired sulfation pathways and immune signaling, especially T cell-mediated responses, may contribute to the pathogenesis mechanism of JIA.

### Estimation of genetic correlations between JIA and other traits

Considering that the pathway analysis showed certain AIDs, such as T1D and AITD, were associated with JIA, we carried out genetic correlation analyses to further investigate the association of JIA with other AIDs. We estimated the genetic correlations between JIA and other traits categorized into two groups, AID-like and non-AID-like traits, at the genome- and transcriptome-wide levels. At the genome-wide level, JIA showed significant positive correlations (FDR_LDSC_ < 0.05) with eight out of the ten (80%) AIDs including systemic lupus erythematosus (SLE) (*r* = 0.66 and FDR_LDSC_ = 1.24 × 10^−08^) and exhibited the most significant correlation with the AID_ALL_ trait (*r* = 0.53 and FDR_LDSC_ = 7.19 × 10^−11^) (Fig. S3A). At the transcriptome-wide level, we found significant genetic overlaps (FDR_RHOGE_ < 0.05) between JIA and seven AID traits including ulcerative colitis (UC), AID_ALL_ trait, Crohn’s disease, rheumatoid arthritis (RA), SLE, primary biliary cirrhosis, and AID_SURE_ trait. JIA was most significantly correlated with UC (*r* = 0.35 and FDR_RHOGE_ = 1.28 × 10^−06^) and had the most positive correlation with T1D (*r* = 0.65 and FDR_RHOGE_ = 6.91 × 10^−02^) (Fig. S3B). In particular, T1D was simultaneously identified to have highly positive correlations with JIA at the genome-(*r* = 0.62 and FDR_LDSC_ = 3.58 × 10^−05^) and transcriptome-wide levels (*r* = 0.65 and FDR_RHOGE_ = 6.91 × 10^−02^). Overall, these results support the existence of shared genetic contributions to JIA and AIDs at the genome- and transcriptome-wide levels.

### TWAS associations were validated by transcriptomic datasets on JIA

To validate TWAS associations are actually related to the gene expression patterns of transcriptomic data, we conducted WGCNA using a microarray dataset from JIA cases and controls (GSE13501)^40^ to identify co-expression modules composed of genes with highly correlated expression patterns (see Methods). A total of 11 co-expression modules were detected and are listed in Table S7 (Fig. S1). First, we validated that the clustering of co-expression modules from the microarray dataset was recapitulated in independent RNA-seq datasets (GSE112057^41^ and GSE79970^42^), respectively (Z-summary score > 2; Fig. S1). Next, we conducted GSEA using the 11 co-expression modules as reference gene sets and TWAS associations of each panel as a ranked gene list. Overall, we observed that TWAS associations from the GTEx visceral omentum adipose panel (normalized enrichment score (NES) = −1.93 and FDR = 2.50 × 10^−02^) and aorta artery panel (NES = −1.93 and FDR = 2.40 × 10^−02^) were significantly enriched in the magenta module (FDR < 0.05; Fig. S2 and Table S8). The TWAS associations from the NTR blood panel and the GTEx skeletal muscle panel were significantly involved in black (NES = 1.84 and FDR = 2.40 × 10^−02^) and yellow (NES = 1.43 and FDR = 4.90 × 10^−02^) modules, respectively (FDR < 0.05). These three modules were highly preserved in both RNA-seq datasets with the Z-summary score > 10 (Fig. S1). We performed functional annotation of the 11 identified modules using gene ontology (GO) terms to identify their biological functions (FDR < 0.05; Table S9)^46, 47^. The significant GO term of the magenta module was “Humoral immune response (GO:0006959)” (FDR = 4.16 × 10^−02^). The black module was significantly associated with “Leukocyte migration (GO:0050900)” (FDR = 7.73 × 10^−5^), while the yellow module was enriched in “Regulation of transcription, DNA-templated (GO:0006355)” (FDR = 4.34 × 10^−8^) and “Transcription, DNA-templated (GO:0006351)” (FDR = 7.11 × 10^−6^). Additionally, significant GO terms of the tan module were “Translational initiation (GO:0006413)” (FDR = 9.78 × 10^−81^) and “rRNA processing (GO:0006364)” (FDR = 8.98 × 10^−71^). Taken together, we observed that TWAS associations were actually enriched in the expression pattern of particular immunological and metabolic gene sets derived from JIA transcriptomic data, suggesting that the TWAS signals may be along with transcriptomic signals for JIA.

### Disease heritability in JIA-relevant tissues and cell types

To better understand how genetic variants affect JIA risks, we aimed to identify the cell types or tissues relevant for the pathogenesis of JIA using LD score regression in specifically expressed genes (LDSC-SEG)^48^. LDSC-SEG tests whether disease heritability is enriched in regions of genes in a specific tissue using stratified LD score regression^87^, analyzing transcriptome or epigenome data together with GWAS. We first applied this analysis to 53 and 152 tissues or cell types from the GTEx project^49^ and Franke lab data^50, 51^, respectively. Consistent with the reason for using connective tissues in the JIA TWAS, this analysis showed that lymphocytes or blood tissues were enriched in JIA (Fig. S4). Additionally, we detected enrichment for the CD4^+^ T cells among 292 immune cell types from ImmGen^52^ in JIA (Fig. S5). To support the results from the expression-based LDSC-SEG analysis, we examined whether JIA-related heritability is enriched in epigenetic markers from the ENCODE projects^54^ and Roadmap Epigenomics^53^. Based on 431 tissue-specific ChIP-seq annotations of six epigenetic marks, we detected an enrichment at the 5% Bonferroni threshold (P < 1.16 × 10^−04^) for only blood tissue (Fig. S6). Notably, the most significant enrichment was observed in T cells across active promoters and gene markers (Fig. 2A) as well as active enhancer markers (Fig. 2B). To validate the Chip-seq results, we used ATAC-seq data which is associated with chromatin accessibility in 13 blood cell types^55^. In line with the Chip-seq results, we found enrichment in T, B, and NK cells after the Bonferroni correction (P < 3.8 × 10^−03^; Fig. 2C).

**Figure 2.**
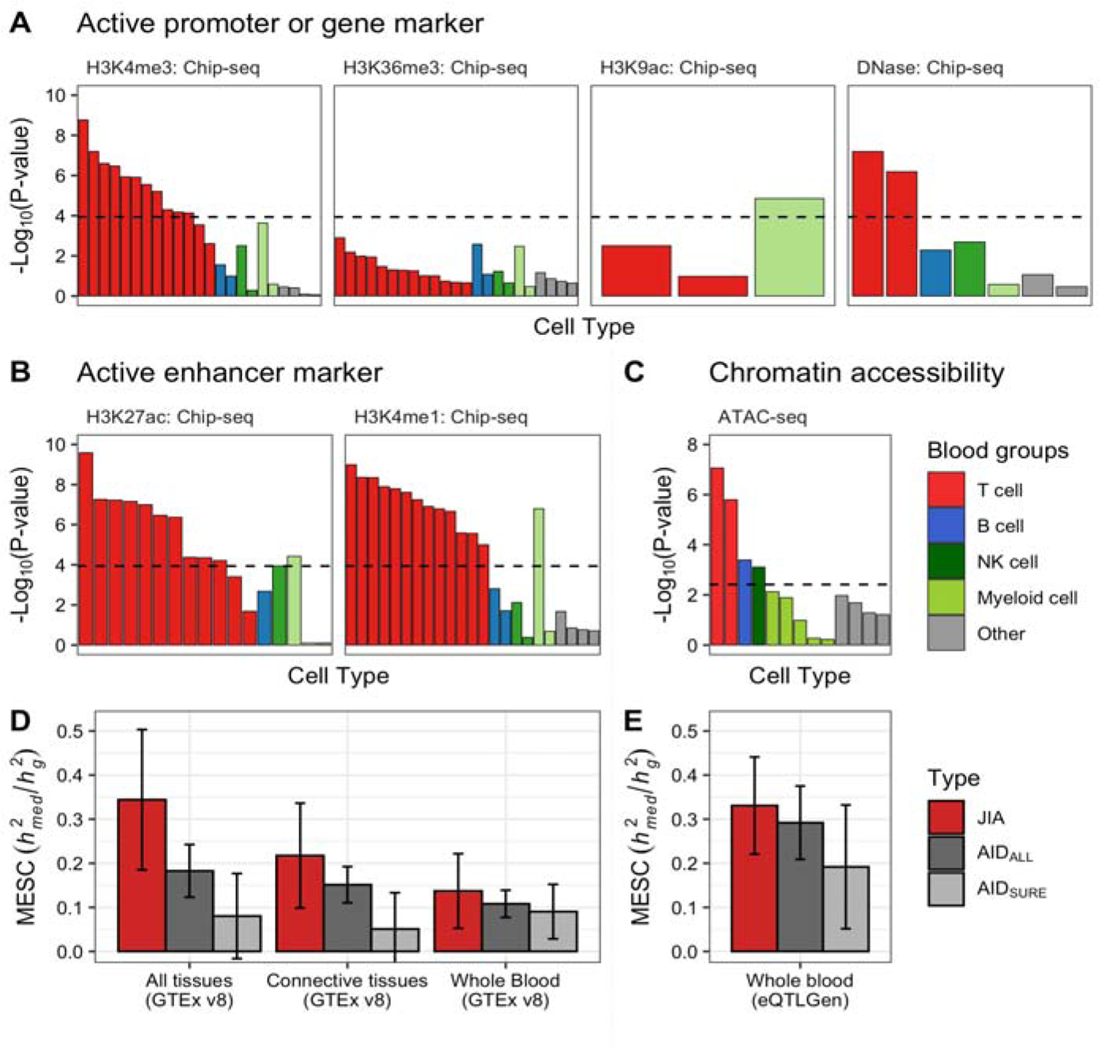
Disease heritability analysis of JIA. (**A-C**) LD score regression in specifically expressed genes (LDSC-SEG) analysis applied to JIA GWAS data using epigenetic markers from blood cell types. (**A**) The enrichment results of LDSC-SEG analysis using active promoter or gene markers. (**B**) The enrichment results of LDSC-SEG analysis using active enhancer markers. The black dotted line represents a significant threshold based on the Bonferroni-corrected P < 1.16 × 10^−04^ (0.05/431). (**C**) The enrichment results of LDSC-SEG analysis using ATAC-seq data. The black dotted line represents a significant threshold based on the Bonferroni-corrected P < 3.85 × 10^−03^ (0.05/13). (**D-E**) Estimation of the proportion of heritability mediated by the gene expression levels (□^2^_□□□_/□^2^_□_) using mediated expression score regression (MESC) for JIA, AID_ALL_, and AID_SURE_. (**D**) The MESC results of all tissues (expression scores from meta-analyses across all 48 GTEx tissues), connective tissues (expression scores from meta-analyses using 12 connective tissues), and whole blood tissue from GTEx v8. (**E**). The MESC results from whole blood tissue from eQTLGen. Error bars indicate jackknife standard errors.

To test whether these enriched tissues and cell types causally mediate JIA risk, we next estimated the proportion of heritability mediated by gene expression levels (□^2^_□□□_/□^2^_□_) in a tissue context using mediated expression score regression (MESC)^57^. When using the all-tissue meta-analyzed expression scores from GTEx, we observed 0.34 of □^2^_□□□_/□^2^_□_ for JIA (standard error (se) = 0.16), which was higher than those from other AIDs (P < 3.37 × 10^−02^; Fig. 2D). We estimated lower □^2^_□□□_/□^2^_□_ from the tissue-group meta-analyzed (i.e., 12 connective tissues) (□^2^_□□□_/□^2^_□_ = 0.217 and se = 0.119) and individual-tissue (i.e., whole blood) expression scores (□^2^_□□□_/□^2^_□_ = 0.137 and se = 0.085) than from all-tissue expression scores in GTEx data (Fig. 2D), consistent with those in previous studies using 42 diseases and complex traits^57^. We identified the highest □^2^_□□□_ value in the EBV-transformed lymphocytes among 48 GTEx tissues (□^2^_□□□_/□^2^_□_ = 0.189 and se = 0.067; Fig. S7), and validated the results by whole blood data from eQTLGen (Fig. 2E)^58^. Together, our findings strongly suggest that the SNP-based heritability of JIA was closely associated with gene expression and active epigenetic markers in blood tissue, especially in T lymphocytes.

### Polygenicity in the genetic architecture of JIA

In complex and polygenic traits, dissecting joint distribution of effect size and MAF is important to understand the genetic architecture and to detect signals of natural selection^88^. Deleterious mutations to fitness are selected against and maintained at a low frequency by negative (purifying) selection^89, 90^. Moreover, the negative selection in autoimmune disease is important because it provides insight into the evolution of the human immune system. We conducted genetic architecture analysis using SBayesS, a recently developed method based on the Bayesian mixed linear model with GWAS summary statistics^88^ to estimate the relationship between SNP effect size and MAF (□^2^_□□□_/□^2^_□_). The SBayesS method also allows us to infer multiple genetic architecture parameters including the SNP-based heritability (h^2^_SBayesS_) and polygenicity (rr) which is the proportion of SNPs with nonzero effects. A negative value of 5 indicates that SNPs with lower MAF are prone to having larger effects, consistent with a model of negative selection. Overall, we estimated 5 = −0.96 (posterior standard error (p.s.e) = 0.10), providing evidence that the genetic variants related to JIA have been under negative selection (Fig. 3A). Excluding SNPs in the HLA region (chr6:28-34Mb) increased the estimate 5 in JIA (Ŝ = −0.49 and p.s.e = 0.2), which suggests that the SNPs in the HLA region contributing towards JIA risk may have been under negative selection. The previous studies showed that the majority of AIDs have genetic relationships with the HLA area, and numerous distinct HLA alleles can predispose people to AIDs^91, 92^. For the SNP-based heritability, the estimate of h^2^ for JIA was 0.47 (p.s.e = 0.03), which was higher when compared with other AIDs (Fig. 3A). We also observed that excluding the HLA region reduced the h^2^ to 0.34 in JIA, which is consistent with previous studies in AIDs^3,93^. Importantly, the polygenicity increased to 8.57% after excluding the HLA region, although the estimated polygenicity (rr) is about 0.09% in JIA.

**Figure 3.**
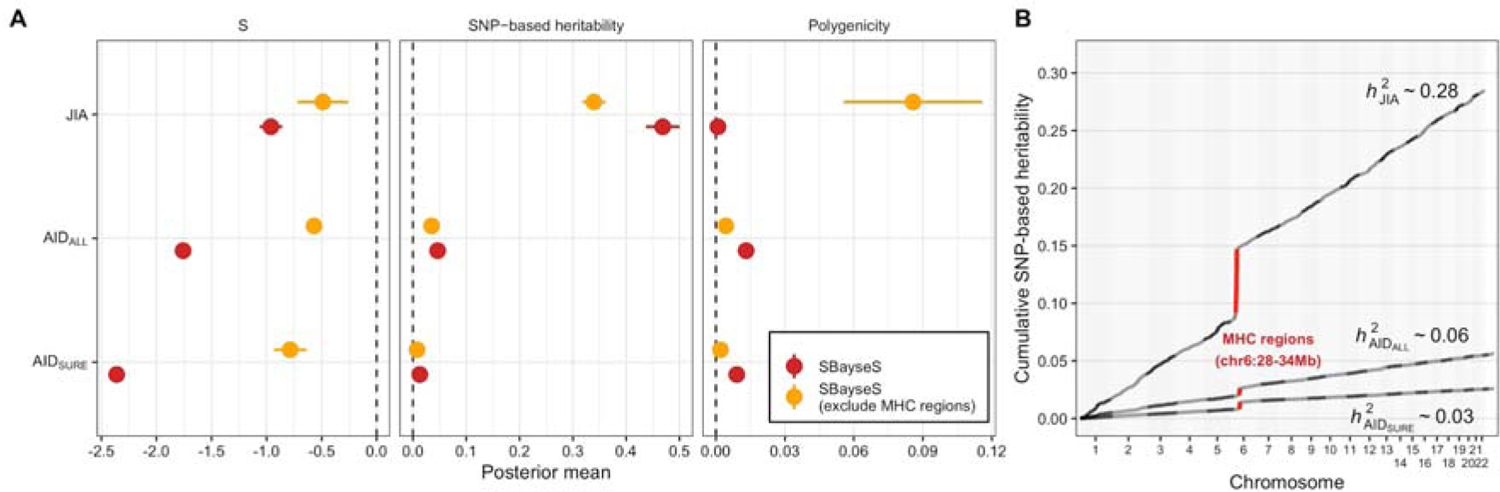
Genetic architecture of JIA. (**A**) Estimation of the three genetic architecture parameters for JIA, AID_ALL_, and AID_SURE_ using Summary-data-based BayesS (SBayesS). The dots and horizontal bars represent the posterior means and standard errors, respectively. The red and orange colors represent SBayesS and SBayesS excluding HLA regions. (**B**) Cumulative local SNP heritability across the genome using heritability estimates from Summary statistics (HESS). Total SNP-based genes are indicated. The red color shows SNP heritability explained by the HLA region.

Moreover, we estimated the SNP-heritability (□^2^_□□□_) using HESS^39^ in each of 1,703 independently partitioned genomic LD blocks across the genome. The results revealed that the total □_□□□□_ was 0.28 in JIA (Fig. 3B), slightly lower than that based on SBayesS (□_□□□□□□□_) (Fig. 3A), and the HLA region significantly explained a total of 17.5% heritability in JIA (Bonferroni-corrected P < 2.94 × 10^−05^; Fig. 3B). In line with the □^2^_□□□_ results, the total □^2^_□□□_ in JIA was higher than the other AIDs. Aside from the HLA region, most of the SNP-based heritability was distributed uniformly across the genome in JIA and other AIDs. Collectively, these results strongly suggest that JIA shows higher polygenicity than other AIDs in both HLA regions and outside of HLA regions, and SNP-based heritability comes from a vastly polygenic background.

### Estimation of the consensus frequencies of HLA allele types and HLA gene expression

The HLA genes in the HLA region on chromosome 6p21 encode several essential proteins in the immune system^94^. Consistent with the SNP-based heritability results in the JIA (Fig. 3B), variants in the HLA regions explain more heritability than many other variants for many diseases^95–97^. Identifying HLA allele type is essential to better understanding the disease etiology, and many tools have been developed for HLA typing using NGS data^42, 60–66^. To explore the association between HLA type diversity and JIA risk, we estimated consensus HLA allele type frequencies at 12 major loci using seven HLA-typing software (see Methods). We focused on the allele types with a significantly different distribution between healthy subjects (*n* = 12) and JIA patients (*n* = 115) (P < 0.05; Table S10). At HLA class I loci, the frequency of A*03:01 was approximately tripled in JIA patients (12.89%) compared with that in healthy subjects (4.22%) (P = 1.10 × 10^−03^; Fig. S8). At HLA class II loci, the frequencies of DPB1*04:01, DQB1*03:02, and DRB1*01:01 were increased by more than two-fold in the JIA group (33.46%, 14.39%, and 10.29%) compared to in the control group (11.21%, 4.23%, and 4.17%), respectively (P_DPB1*04:01_ = 8.99 × 10^−07^, P_DQB1*03:02_ = 7.24 × 10^−04^, and P_DRB1*01:01_ = 1.83 × 10^−02^). Notably, DRB1*04:01 (11.39%) and DRB3*02:01 (11.43%) were only detected in JIA patients. When grouped into three JIA subtypes (oligo-JIA, poly-JIA, and sJIA) (Fig. S9 and Table S11), the frequencies of DRB1*04:01 and DRB3*02:01 were the highest in poly-JIA patients. Additionally, for the transcriptomic analysis of HLA regions, we estimated the locus-specific expression levels of nine HLA genes in JIA patients and healthy controls. The results showed that the expression levels of *HLA-DPA1*, *DPB1*, *DQB1*, and *DRA* were significantly lower in JIA patients than in healthy controls (P < 0.05; Fig. S10). HLA class II is known to be strongly associated with susceptibility to many AIDs including JIA^98–100^. Although various mechanisms could induce the down-regulation of HLA class II gene expression, the lowered expression could result in diminished tolerance induction of self-reactive T cells, leading to AIDs in the end^101^.

In the HLA region, haplotypes are specific to an individual ancestral population due to the population-specific positive selection^102^. We calculated the consensus frequencies of HLA types of JIA in African and European ancestry to investigate whether heterogeneity is derived from different ancestry in HLA allele types for JIA. We focused on the allele types with significantly different distributions between healthy controls (*n* = 12), African JIA (*n* = 34), and European JIA patients (*n* = 81) (P < 0.05; Table S12). At HLA class I loci, B*45:01 was not detected in healthy controls and was more frequent in African JIA patients (11.18%) than European JIA patients (0.62%) (P = 8.33 × 10^−22^; Fig. 4A). B*40:01, which was not observed in African patients, had higher frequencies in European patients (10.65%) than in healthy controls (5.36%) (P = 4.90 × 10^−02^). At HLA class II loci, the frequencies of DQA1*01:01, and DQA1*03:01 were increased by more than four-fold in the European JIA patients (13.68% and 12.84%) compared to in the African JIA patients (3.05% and 1.78%), respectively (P_DQA1*01:01_ = 9.65 × 10^−08^, P_DQA1*03:01_ = 2.56 × 10^−09^; Fig. 4B). The frequencies of DRB1*04:01 observed only in JIA patients, was higher in European patients (15.10%) than African patients (2.49%) (P = 1.02 × 10^−09^; Fig. 4B). Collectively, our results showed that HLA allele types had significantly imbalanced distributions between the JIA and control groups as well as between African and European ancestry, which suggests that they may be involved in JIA etiologies.

**Figure 4.**
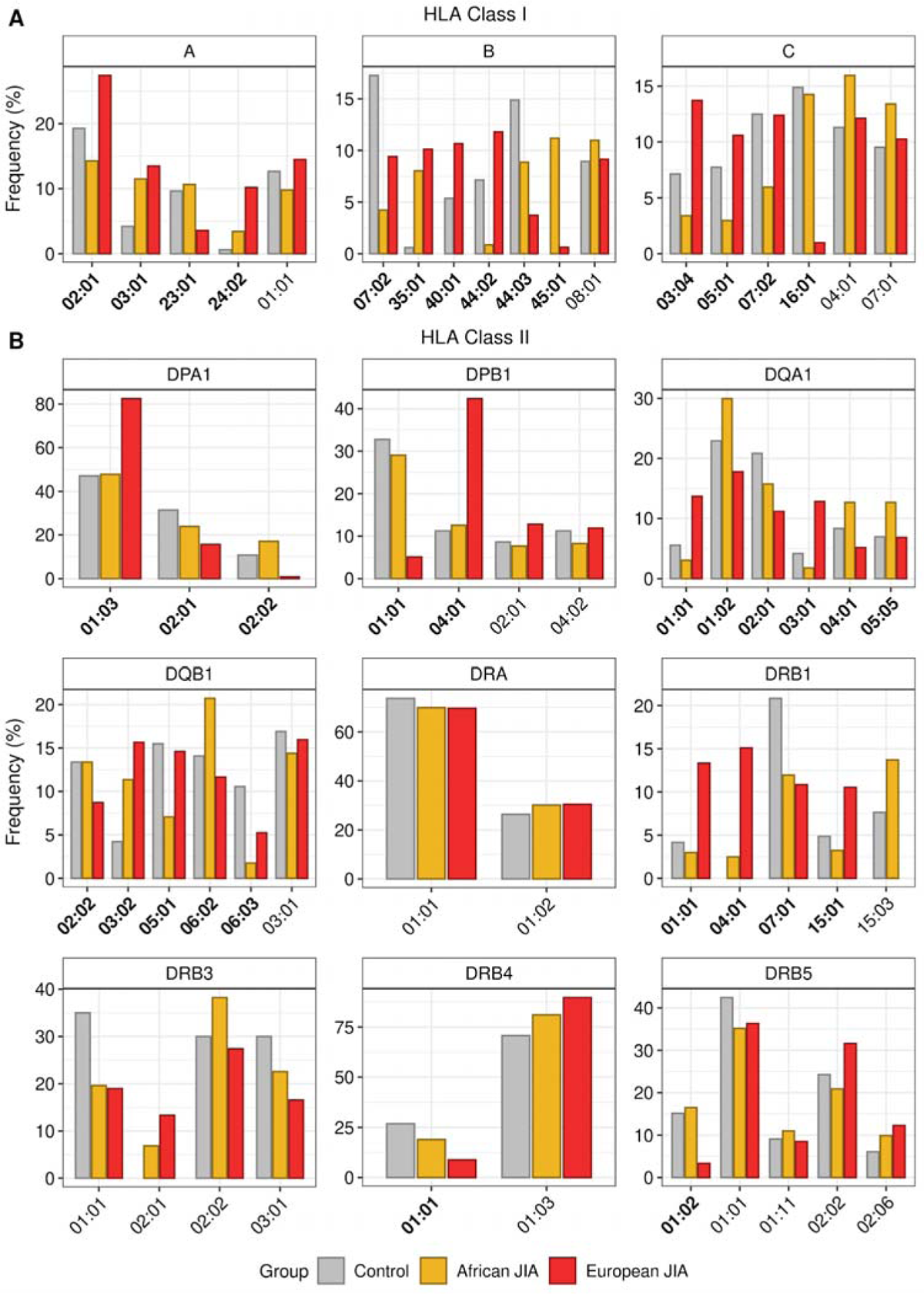
Consensus frequencies of HLA allele types in healthy controls, African JIA patients, and European JIA patients. (**A**) Bar plots showing the consensus frequencies of HLA allele types at HLA class I loci. (**B**) Bar plots showing the consensus frequencies of HLA allele types at HLA class II loci. The x- and y-axis indicate the names and frequencies of HLA allele types, respectively. The names of HLA types having significantly different distributions between healthy controls, African JIA patients, and European JIA patients were represented in bold (P < 0.05).

### The T cell receptor repertoire reveals clonal relationships between different subpopulations

Along with HLA molecules, antigen-experienced memory T cells have been implicated as critical drivers of autoimmune inflammation^103–106^. Consistent with these previous studies, we suggested that T cells were associated with JIA etiology using TWAS and JIA SNP-heritability analysis. Therefore, we hypothesize that the TCR repertoire may be involved in JIA etiology because the TCRs mediate the recognition of HLA and provide critical insights into the adaptive immune response in health and disease^107^. To dissect whether the TCR repertoire is related to JIA, we estimated the locus-specific alpha diversities of TCR CDR3 reads using the total number of distinct clonotypes and their relative frequencies at the bulk RNA-seq level (GSE112057^41^). We observed alpha diversities of TCR α and β were significantly decreased in JIA patients compared with controls (P_TCRα_ = 9.60 × 10^−04^ and P_TCRβ_ = 1.36 × 10^−02^), which indicates that the clonotypic diversities of TCRs were reduced in the JIA group (Fig. 5A). When JIA was grouped into three subtypes (oligo-JIA, poly-JIA, and sJIA), the patterns of alpha diversity in all subtypes were consistent with those in the JIA group (Fig. S11 and Table S13). These results suggest that a few clonotypes positively affecting the development of JIA may be dominant in the immune repertoire of JIA patients.

**Figure 5.**
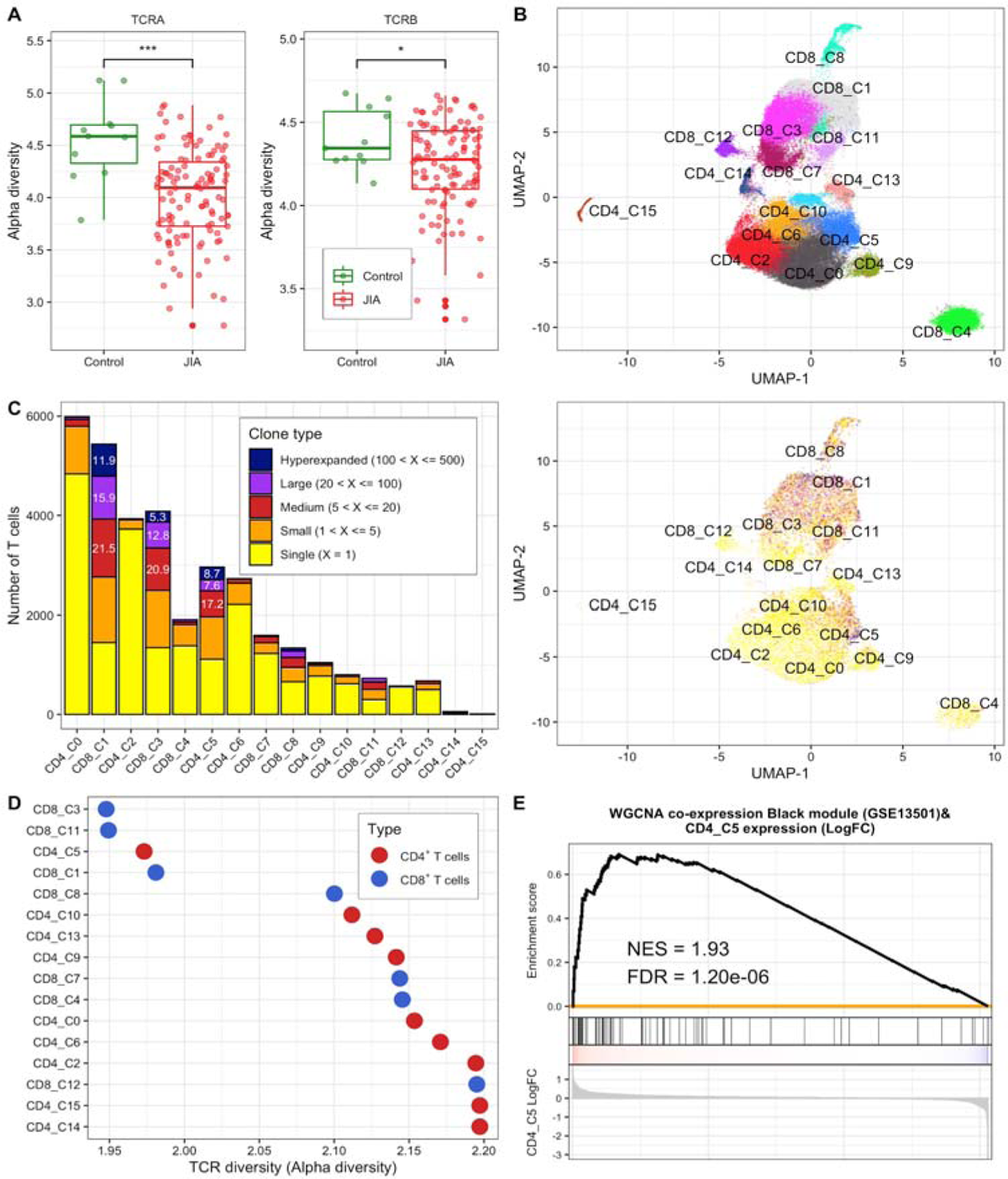
TCR diversities in JIA. (**A**) Box plots showing the alpha diversities of TCR α and β in healthy control and JIA groups. The y-axis indicates alpha diversity representing the clonotypic diversity of specific TCR locus. Green and red dots indicate samples of healthy controls and JIA patients, respectively. Asterisks denote the significance levels of differences between the clonotypic diversities of TCRs within healthy controls and that within JIA patients. *, P < 0.05; **, P < 0.01; ***, P < 0.001. (**B**) A Uniform Manifold Approximation and Projection (UMAP) plot of 67,235 T cells from seven JIA patients showing 16 major clusters (nine for 34,605 CD4^+^ and seven for 32,630 CD8^+^ T cells). Each dot represents an individual T cell and color indicates cluster origin. (**C**) Left. A bar plot showing the number of single T cells and frequencies of unique and expanded T cell clones in each cluster. The inset numbers indicate the proportion of the cell type in the cluster. Right. UMAP plot showing the clusters with the clone types. The colors correspond to the number of clones (i.e. clonal abundance). (**D**) A scatter plot showing the alpha diversity of TCR in each cluster. The red and blue colors denote CD4^+^ and CD8^+^ T cells, respectively. (**E**) Module enrichment analysis between expression levels of scRNA-seq CD4_C5 cluster and black co-expression gene set derived from JIA case-control expression data (GSE13501). The gene list of the black modules is in Table S9.

Next, we investigated whether different T cell populations showed differences in TCR diversity at the single-cell level. Single-cell TCR sequencing can accurately measure the diversity of T cell populations, which is critical for understanding the complexity of the immune response by providing paired TCR α and β information^108^. We used scRNA-seq data (SRP288574^72^) derived from single CD4^+^CD45RO^+^CD25^−^ (CD4^+^) and CD8^+^CD45RO^+^ (CD8^+^) T cells in SF and PB tissues of seven oligo-JIA patients. After a series of quality control filters (see Methods), nine CD4^+^ and seven CD8^+^ T cell clusters were identified (Fig. S12, S13, and 5B), and each cluster represented a distinct distribution of clonotypes (Fig. 5C). Among 67,235 single T cells, 33,855 cells (50.3%) had at least one pair of full-length TCR α and β chains. In addition, 13,113 of the 33,855 cells (38.7%) expressed a full-length α-β chain pair detected at least twice. Expanded clonotype cells were most prevalent in two CD8^+^ clusters (CD8_C1 and CD8_C3) and one CD4^+^ cluster (CD4_C5) (Fig. 5C). Compared with the other clusters, the CD8_C1 cluster had higher expression of several markers of recently activated effector memory or effector T cells (designated as CD8^+^ *GZMH^+^* T_EMRA_ cells) and the CD8_C3 cluster had higher expression of several markers of effector memory T cells (designated as CD8^+^ *GZMK^+^* T_EM_ cells) (Table S14). The CD4_C5 cluster showed higher expression of *CXCL13* and *BHLHE40* (designated as *CXCL13*^+^*BHLHE40*^+^ T_H_ cells) (Fig. S14 and Table S14). Each cluster of the CD8^+^ *GZMH^+^* T_EMRA_, CD8^+^ *GZMK^+^* T_EM_, and *CXCL13*^+^*BHLHE40*^+^ T_H_ cells was observed to have a substantially lower alpha diversity value than most other clusters (Fig. 5D). Additionally, the single T cell analysis by RNA-seq and TCR tracking expansion (STARTRAC-expa) index, which quantitatively describes tissue clonal expansion, revealed the CD8^+^ *GZMH^+^* T_EMRA_, CD8^+^ *GZMK^+^* T_EM_, and *CXCL13*^+^*BHLHE40*^+^ T_H_ cells as the clusters with the highest degree of clonal expansion for each cell type (Fig. S15).

To emphasize disease-specific memory T cell clusters, we conducted GSEA using the co-expression modules from case-control JIA expression data as reference gene sets (Fig. S16). The results showed that only the *CXCL13*^+^*BHLHE40*^+^ T_H_ cells were positively enriched in the black module gene set (NES = 1.93 and FDR = 1.20 × 10^−6^; Fig. 5E). Notably, the black module genes, associated with leukocyte migration essential for inflammation and innate immunity^109^ (Table S9), were positively enriched in TWAS associations from NTR blood tissue (Fig. S2C). In summary, we identified clonally expanded T cell subpopulations in JIA patients. The *CXCL13*^+^*BHLHE40*^+^ T_H_ cells were significantly related to case-control JIA expression data, suggesting that the cells might be potential therapeutic targets for JIA.

## Discussion

Using a TWAS/PWAS to identify biologically interpretable susceptibility genes/proteins, we detected 19 TWAS genes and two PWAS proteins significantly associated with JIA risks (Fig. 1). Since trait-associated genes/proteins identified by TWAS/PWAS do not fully elucidate the disease’s causality^110^, we performed a fine-mapping analysis to prioritize putatively causal genes/proteins for JIA. We found that *MAGI3*, *DCLRE1B*, *NFATC2IP*, IL27, and ERAP2 were responsible for JIA risks and are implicated in the immune system. *MAGI3* encodes the PDZ proteins involved in T cell homeostasis and mediates the suppression of the PI3K/Akt pathway^111, 112^. The down-regulation of *MAGI3* may lead to an up-regulation of the PI3K/Akt pathway, which, in turn, down-regulates the differentiation of T cells toward Treg^10, 112^. *DCLRE1B* was reportedly implicated in an inherited bone marrow failure syndrome associated with immune deficiency^113^. *NFATC2IP* regulates the nuclear factor of activated T cells (NFAT)-driven transcription of specific cytokine genes, including *IL4* in T-helper 2 cells^114^. Considering that IL4 production is affected by IL27^115^, *NFATC2IP* may be linked to the inflammatory cytokine pathways identified by the TWAS-GSEA (Table 1). ERAP2 reportedly has crucial roles in immunomodulating immune responses^116^. While our TWAS genes, *MAGI3* and *NFATC2IP*, were mentioned in previous JIA TWAS studies^81, 117, 118^, we could better understand the genetic contribution of risk genes to JIA pathogenesis by conducting a PWAS together with fine-mapping and TWAS-based pathway enrichment analysis. A proteome-level analysis can provide more relevant biological information, capture alternative splicing, and post-translational modifications about disease mechanisms.

In genetic architecture analysis, SNP-based heritability was spread uniformly throughout the genome aside from a modest fraction in the HLA regions (about 18%) (Fig. 3). The results can be explained by an “omnigenic model”. The omnigenic model is a theoretical framework in genetics and genomics that proposes that most traits, diseases, and other complex phenotypes are influenced by the combined effects of a large number of genes, rather than being driven by just a few “core” genes^119, 120^.

Alongside the TWAS/PWAS and genetic architecture analysis, we conducted HLA-typing analyses as per the disease state and ancestry using seven HLA-typing software. We suggested DRB1*04:01 and DRB3*02:01 showing >10% consensus frequencies in only JIA patients as risk alleles for JIA. In line with our result, it was reported that DRB1*01 and DRB1*04 might be implicated in the genetic predisposition of rheumatoid factor+ JIA and that DRB1*04 was confirmed to be involved in sJIA^121^. In addition, DRB1*04:01 and DQB1*03:02 may have a shared contribution to JIA and T1D since specific interactions between DRB1*03:01-DQB1*02:01/DRB1*04:01-DQB1*03:02 genotypes were previously described to increase T1D risks^122^. As the distribution of HLA alleles varies among different ancestries, multi-population needs to be considered an important factor in studying the associations between HLA allele types and disease risks^123^. Through HLA-typing analysis utilizing JIA patients’ multi-ancestry information, we identified ancestry-specific risk alleles in both HLA class I (B*40:01 and B*45:01) and HLA class II (DQA1*01:01, DQA1*03:01, and DRB1*04:01) (Fig. 4). Even though some HLA types were previously reported as risk alleles for JIA in specific populations, few HLA studies have compared risk alleles for JIA between different ancestral groups^124, 125^.

A recent study showed that there is a hypothesis that HLA risk alleles may affect the risks of autoimmunity by influencing thymic T cell selection^126^. T cells with receptors that recognize self-HLA molecules and interact with foreign antigens are selected during T cell development in the thymus^127^. In AIDs, self-antigens may induce immune responses by self-reactive T cells similar to how foreign antigens trigger immune responses^128^. As the proliferation of antigen-specific lymphocytes is induced by immune responses, we believe that the reduced alpha diversities of TCRs within JIA patients may be attributed to the selective increase of a few different self-reactive clonotypes at the levels of individual patients (Fig. 5). In fact, circulating CD4^+^ T cells replicating the phenotypical signature of T lymphocytes infiltrating the inflamed synovium were increased in patients with JIA^129^. Previous clinical studies also reported that the alpha diversity of the TCR repertoire was significantly reduced in patients with RA or SLE that are also AIDs^130–132^. At the single-cell level, our results showed that clonally expanded T cell subpopulations in JIA patients, especially *CXCL13*^+^*BHLHE40*^+^ T_H_ cells, were significantly involved in JIA risks (Fig. 5). The *PD-1*^+^*TOX*^+^*BHLHE40*^+^ population of CD4^+^ T cells was reported to presumably support extrafollicular B cell activation by secreting IL21 and CXCL13 in JIA^72^. In addition, it was reported that CXCL13-producing CD4^+^ T_H_ cells induced in RA synovium may be involved in the recruitment of B cells and circulating follicular helper T cells at inflammation sites^133^.

Although our study successfully identified susceptibility genes/proteins for JIA by TWAS/PWAS, functional studies are needed to clarify the exact genetic effects derived from the genes/proteins. We excepted genes from the HLA region due to its structural diversity and long-range LD in the TWAS/PWAS; however, the GWAS trait-associations have been more reported in the HLA region than in any other locus^11^. To compensate for this situation, we identified the consensus HLA allele types using seven different HLA-typing tools with RNA-seq data. The genetic architecture analysis, especially for SNP-based heritability, requires additional confirmation because we estimated the heritability using GWAS summary statistics data with reference LD dataset. In addition, the publicly available data we used had some limitations. While GWAS summary statistics data for JIA comprising various subtypes was used because there were very few publicly available GWAS data for each JIA subtype, the scRNA-seq dataset was derived solely from oligo-JIA patients. Since the RNA-seq dataset for HLA typing did not contain ancestral information on healthy subjects, the ancestry-specific risks of HLA alleles need to be further confirmed in the control group. Despite these limitations, we believe that this study sheds new light on the pathogenesis of JIA and provides a strong foundation for future mechanistic studies aimed at uncovering the molecular drivers of JIA.

## Data Availability

All data produced in the present work are contained in the manuscript.

## Acknowledgement

This work was a part of the fulfillment of the requirements for Daeun Kim’s M.S. degree.

## Author Contribution

Conceptualization, D.K. and J.J.; methodology, D.K., N.M., S.M. and J.J.; software, D.K., J.J., and J.S.; validation, D.K. and J.J.; formal analysis, D.K. and J.J.; investigation, D.K. and J.J.; data curation, D.K., J.J., and J.S.; writing—original draft preparation, D.K. and J.J.; writing—review and editing, J.J., C.W.A. and W.J.; visualization, D.K., J.J., and J.S.; supervision, J.J. and W.J.; project administration, J.J. and W.J.; All authors have read and agreed to the published version of the manuscript.

## URLs

FUSION: https://github.com/gusevlab/fusion_twas

FOCUS: https://github.com/bogdanlab/focus

TWAS-GSEA v1.2: https://github.com/opain/TWAS-GSEA

MAGMA v1.07: https://ctg.cncr.nl/software/magma

MSigDB v7.2: https://www.gsea-msigdb.org/gsea/msigdb/

RHOGE: https://github.com/bogdanlab/RHOGE

TWAS-hub: http://twas-hub.org/

ARIC: http://nilanjanchatterjeelab.org/pwas/

LD-hub: http://ldsc.broadinstitute.org/

UKBB: https://alkesgroup.broadinstitute.org/UKBB/

fgsea v1.19.2: https://github.com/ctlab/fgsea

DAVID: https://david.ncifcrf.gov/

WGCNA: https://horvath.genetics.ucla.edu/html/CoexpressionNetwork/Rpackages/WGCNA/

LDSC v1.0.1: https://github.com/bulik/ldsc

GCTB v2.03 beta: https://cnsgenomics.com/software/gctb/

MESC: https://github.com/douglasyao/mesc

GTEx: https://gtexportal.org/home/

eQTLGen: https://www.eqtlgen.org/

HESS v0.5.3 beta.: https://huwenboshi.github.io/hess/

STAR v2.5.3a: https://github.com/alexdobin/STAR

IMGT/HLA database v3.10.0: https://www.ebi.ac.uk/ipd/imgt/hla/

seq2HLA v2.3: https://github.com/TRON-Bioinformatics/seq2HLA

arcasHLA v0.2.0: https://github.com/RabadanLab/arcasHLA

HLAforest: https://github.com/FNaveed786/hlaforest

HLA-VBSeq v2: http://nagasakilab.csml.org/hla/

OptiType v1.3.3: https://github.com/FRED-2/OptiType

PHLAT v1.0: https://sites.google.com/site/phlatfortype/download

HLA-HD v1.2.1: https://www.genome.med.kyoto-u.ac.jp/HLA-HD/

ROP: https://github.com/smangul1/rop

ImReP: https://github.com/mandricigor/imrep

cellranger v6.1.2 and human (GRCh38) References v 2020-A: https://support.10xgenomics.com/single-cell-gene-expression/software/downloads/latest

Seurat v4.1.0: https://satijalab.org/seurat/

harmony v0.1.0: https://github.com/immunogenomics/harmony

