## Supplementary Figures for "Large-scale Integrative Analysis of Juvenile Idiopathic Arthritis for New Insight into Its Pathogenesis"

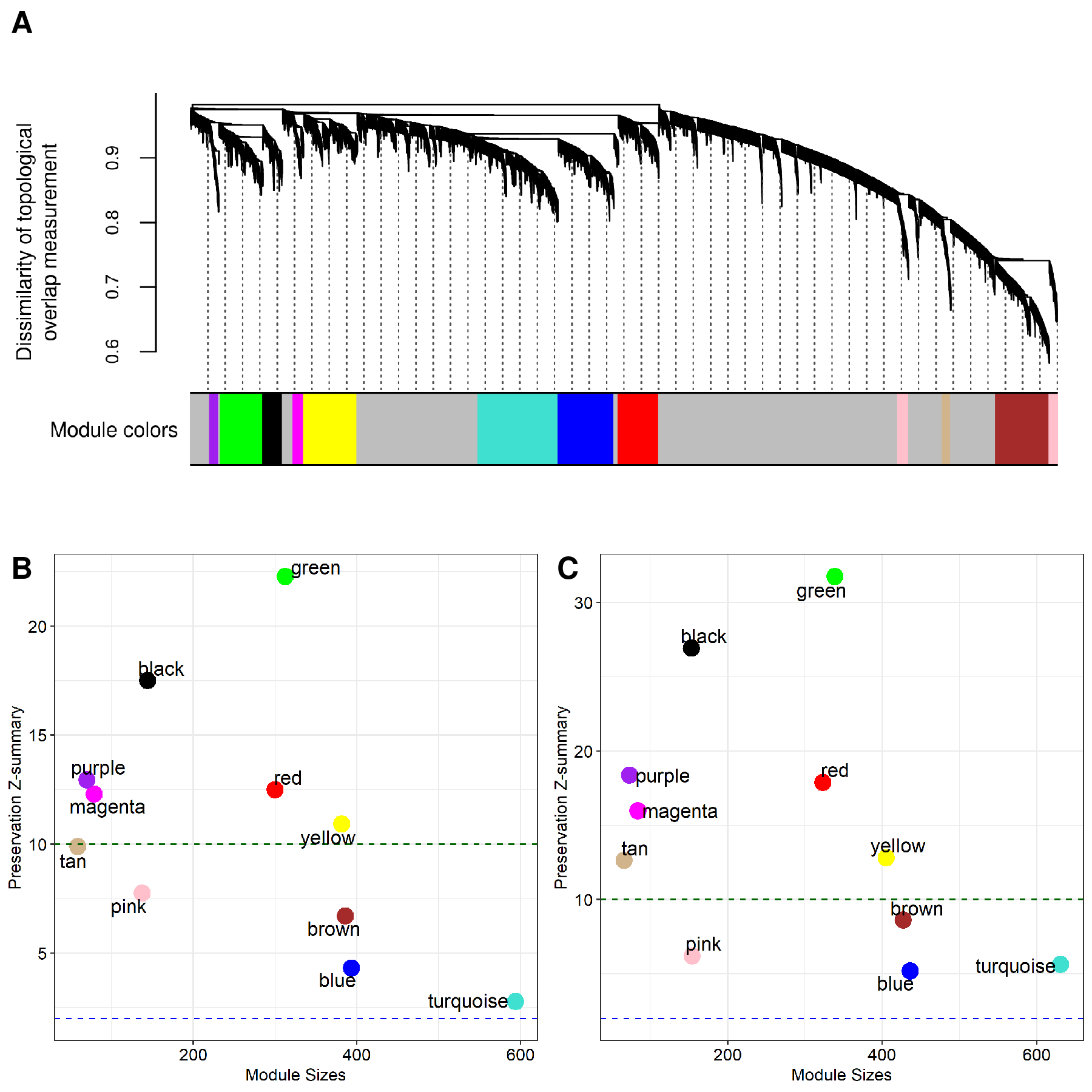

**Supplementary Figure S1. Identification of co-expression modules using GSE13501 and WGCNA.** (**A**) A dendrogram showing co-expression modules based on the dissimilarity of topological overlap measurement. Color bars represent the randomly assigned colors for the module names. The orders of module colors are purple, green, black, magenta, yellow, turquoise, blue, red, pink, tan, and brown. (**B**) The result of module preservation analysis using GSE13501 as a reference set and GSE112057 as a test set. (**C**) The result of module preservation analysis using GSE13501 as a reference set and GSE79970 as a test set. The green and blue dotted lines indicate the thresholds of significance, respectively (Z-summary score > 2 and Z-summary score > 10).

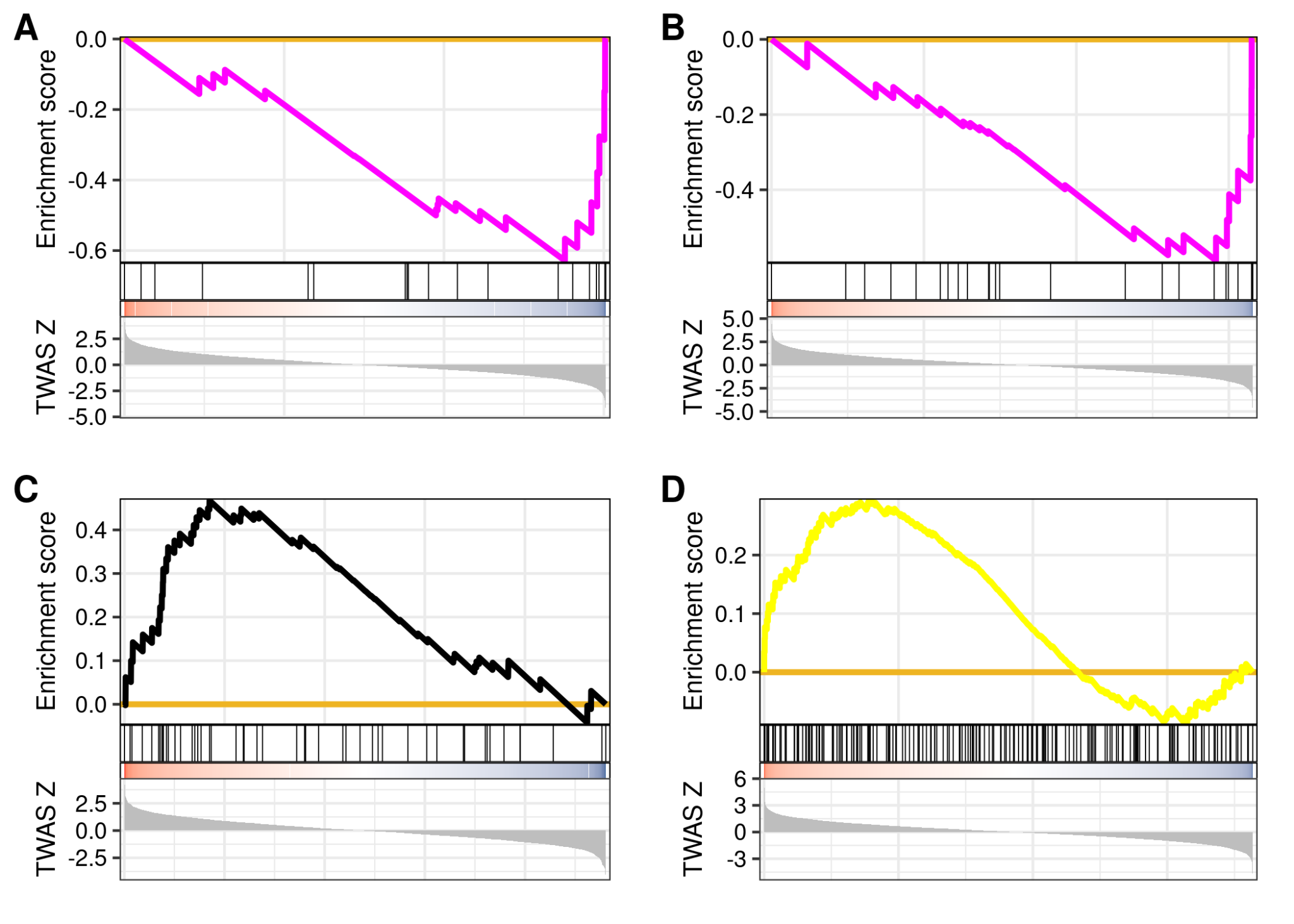

**Supplementary Figure S2. Functional annotation of co-expression modules enriched with TWAS associations.** (**A**) A GSEA plot using a ranked gene list from GTEx: Adipose Visceral Omentum and magenta module (normalized enrichment score (NES) = −1.93 and FDR = 0.025) (**B**) A GSEA plot using a ranked gene list from GTEx: Artery Aorta and magenta module (NES = −1.93, and FDR = 0.024). (**C**) A GSEA plot using a ranked gene list from NTR: Blood and black module (NES = 1.84, and FDR = 0.024). (**D**) A GSEA plot using a ranked gene list from GTEx: Muscle Skeletal and yellow module (NES = 1.43, and FDR = 0.049).

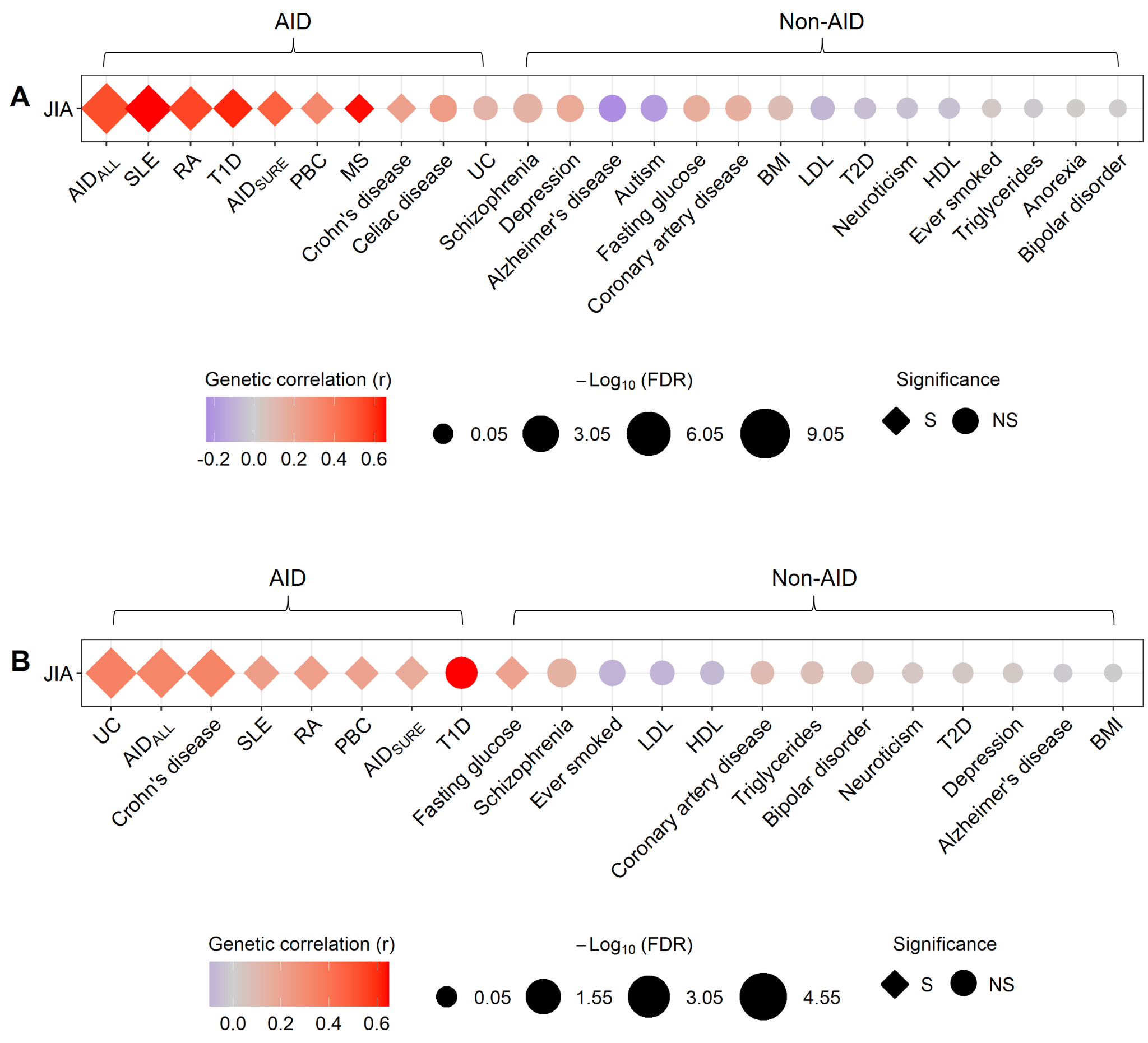

**Supplementary Figure S3. Genetic correlations between JIA and other traits at the genome- and transcriptome-wide levels.** AID-like and non-AID-like traits were compared with JIA. The range of genetic correlation coefficient and FDR values are represented by a color bar and symbol size, respectively. The shape of the symbol indicates the significance of the corresponding correlation. (**A**) Genetic correlations between JIA and other traits at the genome-wide level. (**B**) Genetic correlations between JIA and other traits at the transcriptome-wide level, based on the TWAS results. S stands for significant and NS for non-significant.

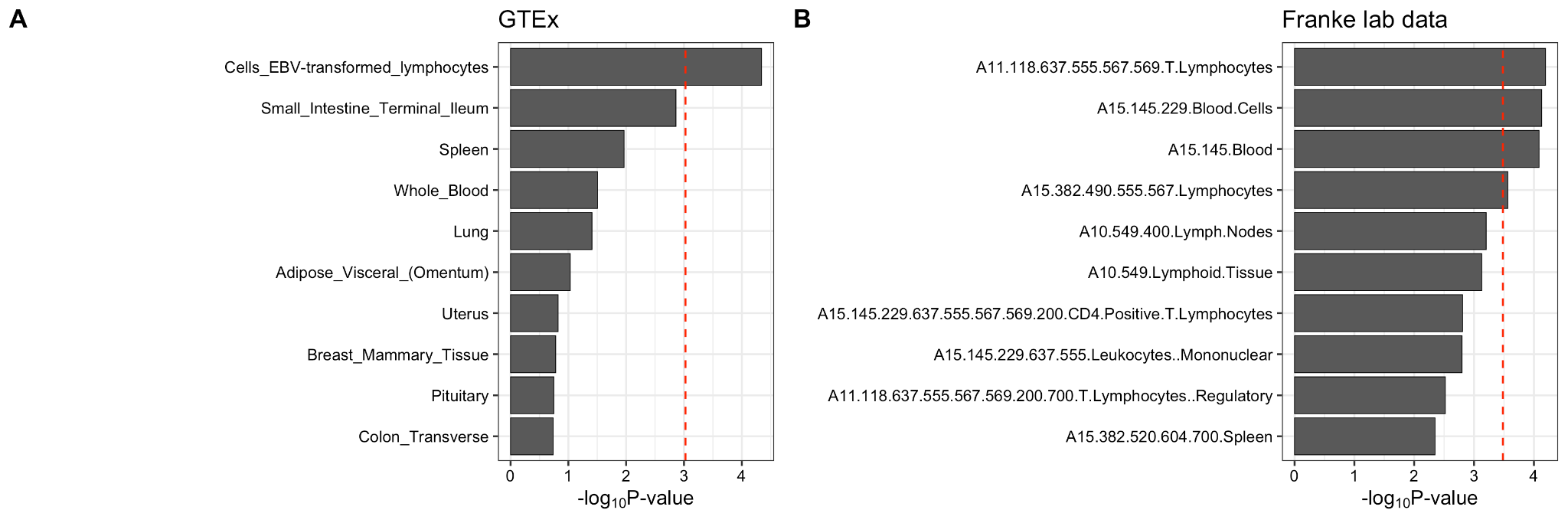

**Supplementary Figure S4. Heritability enrichment analysis using tissue or cell type expression.** Linkage disequilibrium (LD) score regression in specifically expressed genes (LD-SEG) analysis applied to JIA GWAS data. (**A**) The enrichment results of LD-SEG analysis using GTEx data. (**B**) The enrichment results of LD-SEG analysis using Franke lab data. The red dotted lines represent a significant threshold based on the Bonferroni correction. P-values of 9.43 × 10^−04^ (0.05/53) and 3.28 × 10^−04^ (0.05/152) are the significant thresholds of the results from GTEx and Franke lab data, respectively.

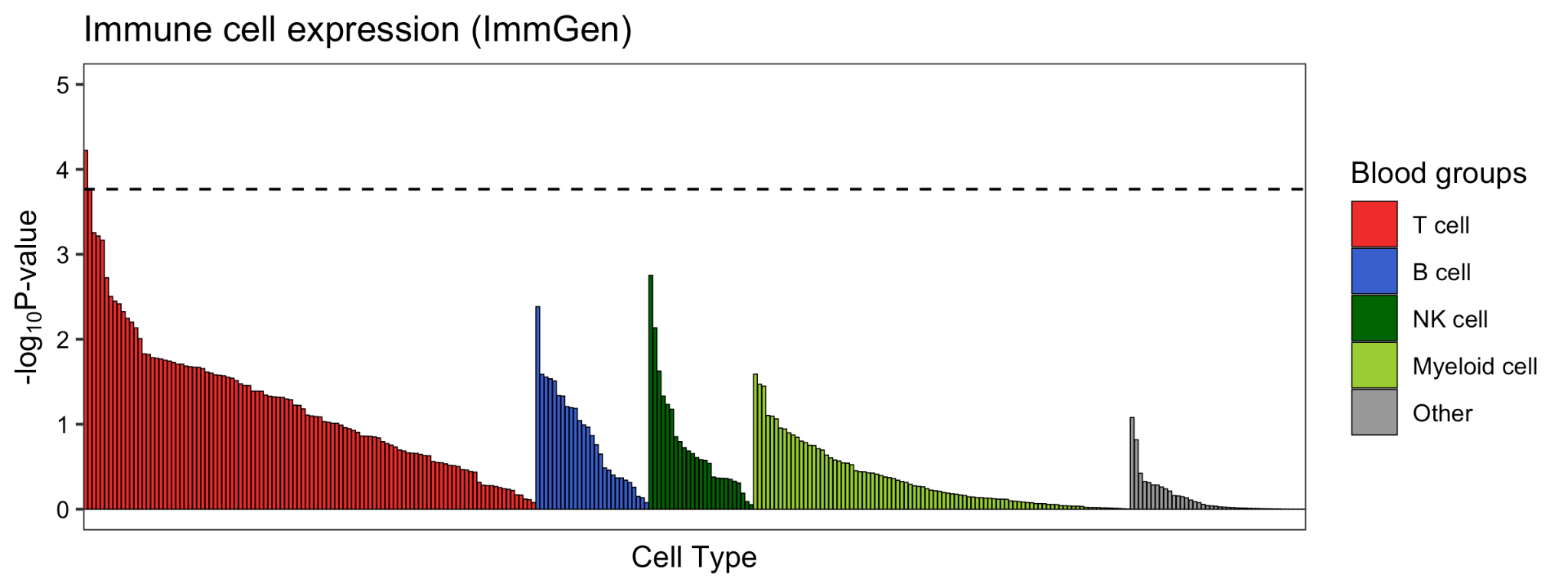

**Supplementary Figure S5. Heritability enrichment analysis using immune cell type expression.** The enrichment results of LD-SEG analysis using ImmGen datasets. The black dotted line represents a significant threshold based on the Bonferroni-corrected P < 1.71 × 10^−04^ (0.05/292).

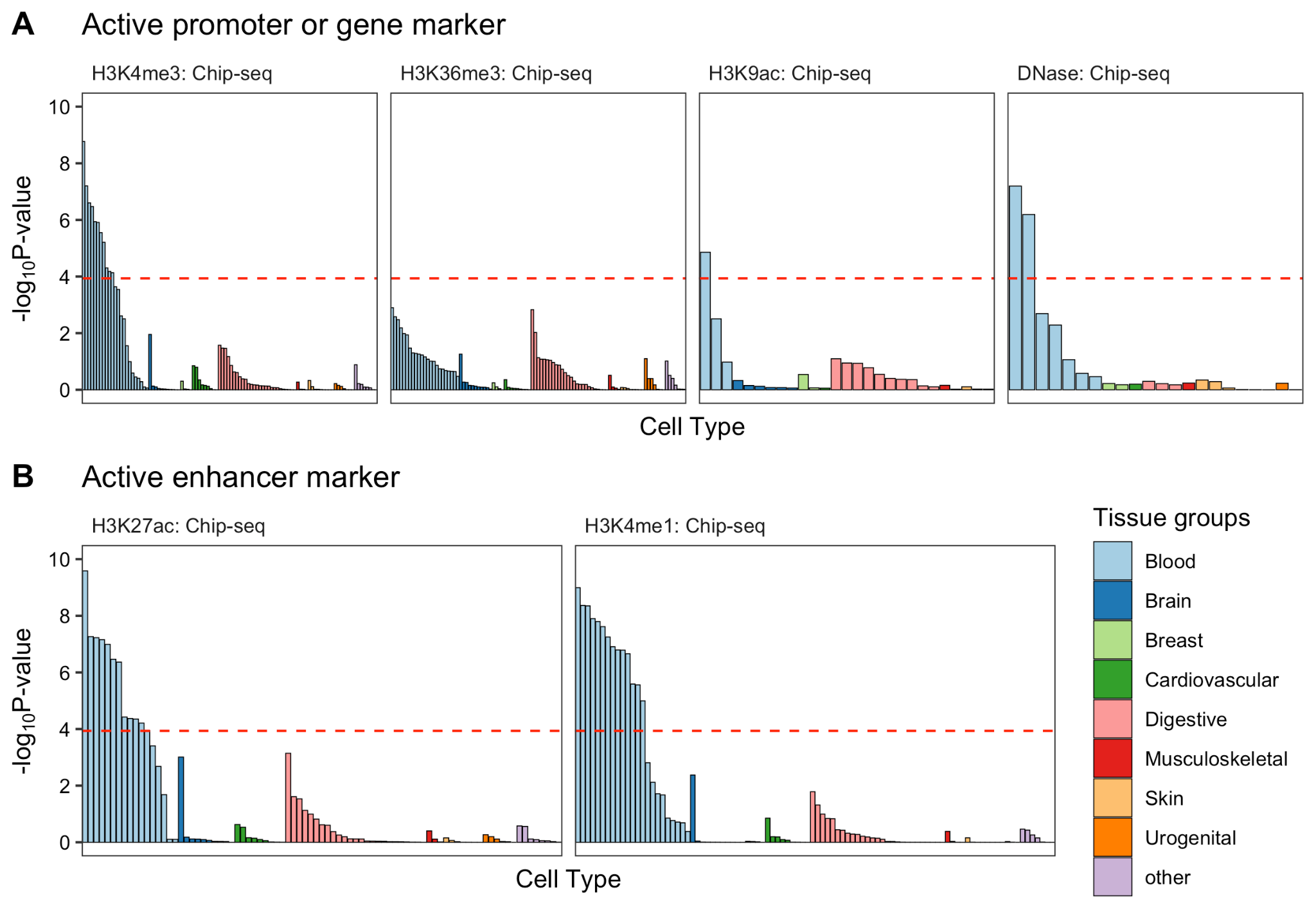

**Supplementary Figure S6. Heritability enrichment analysis using epigenetic markers.** The enrichment results of LD-SEG analysis using epigenetic markers of different tissue types. The red dotted lines represent a significant threshold based on the Bonferroni-corrected P-value < 1.16 × 10^−04^ (0.05/431).

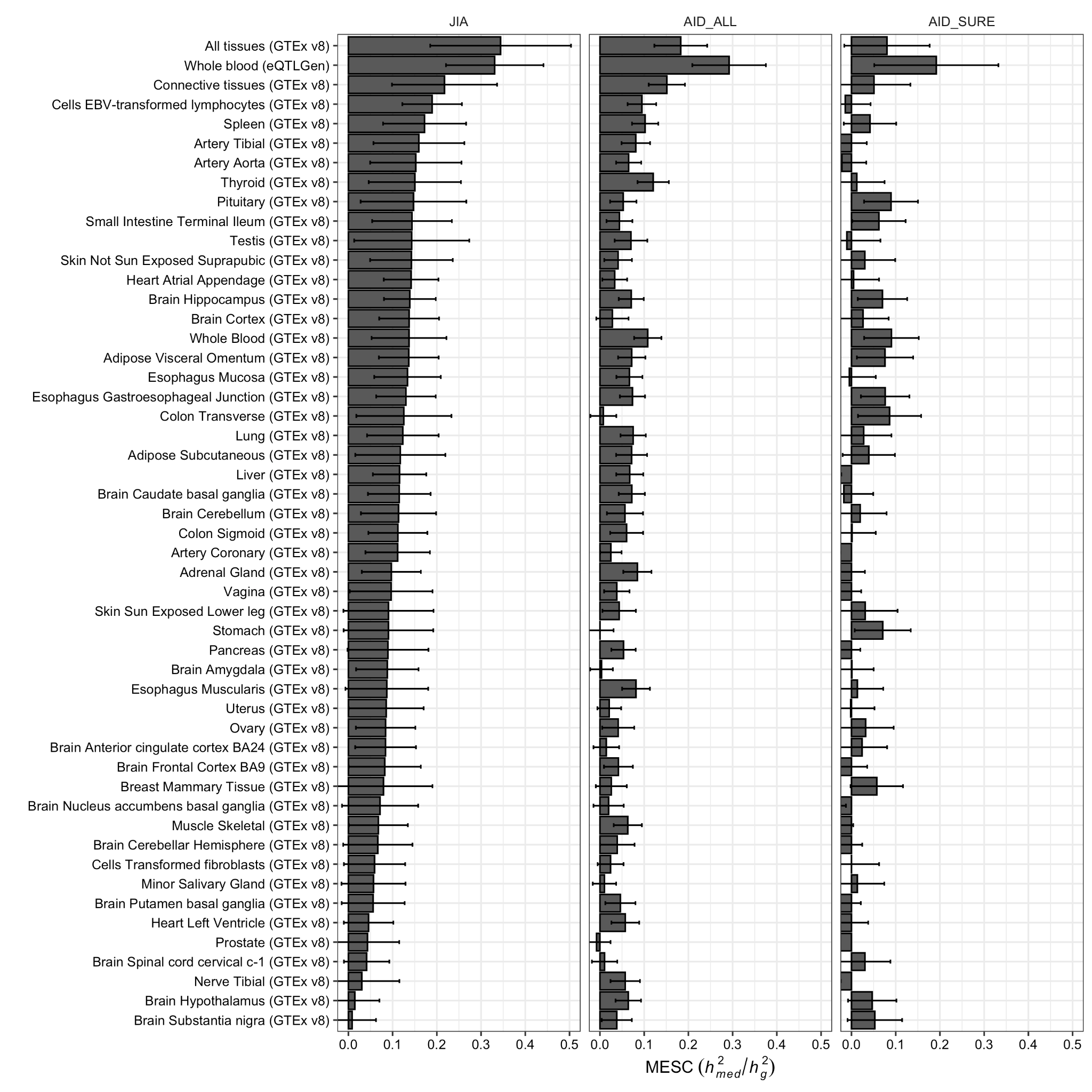

**Supplementary Figure S7.**  **Estimation of the proportion of heritability mediated by the gene expression levels (**$\boldsymbol{h}_{\boldsymbol{med}}^{\boldsymbol{2}}\boldsymbol{/}\boldsymbol{h}_{\boldsymbol{g}}^{\boldsymbol{2}}$**).** The bar plots show the results of mediated expression score regression (MESC) using GTEx v8 and eQTLGen data. Error bars indicate jackknife standard errors.

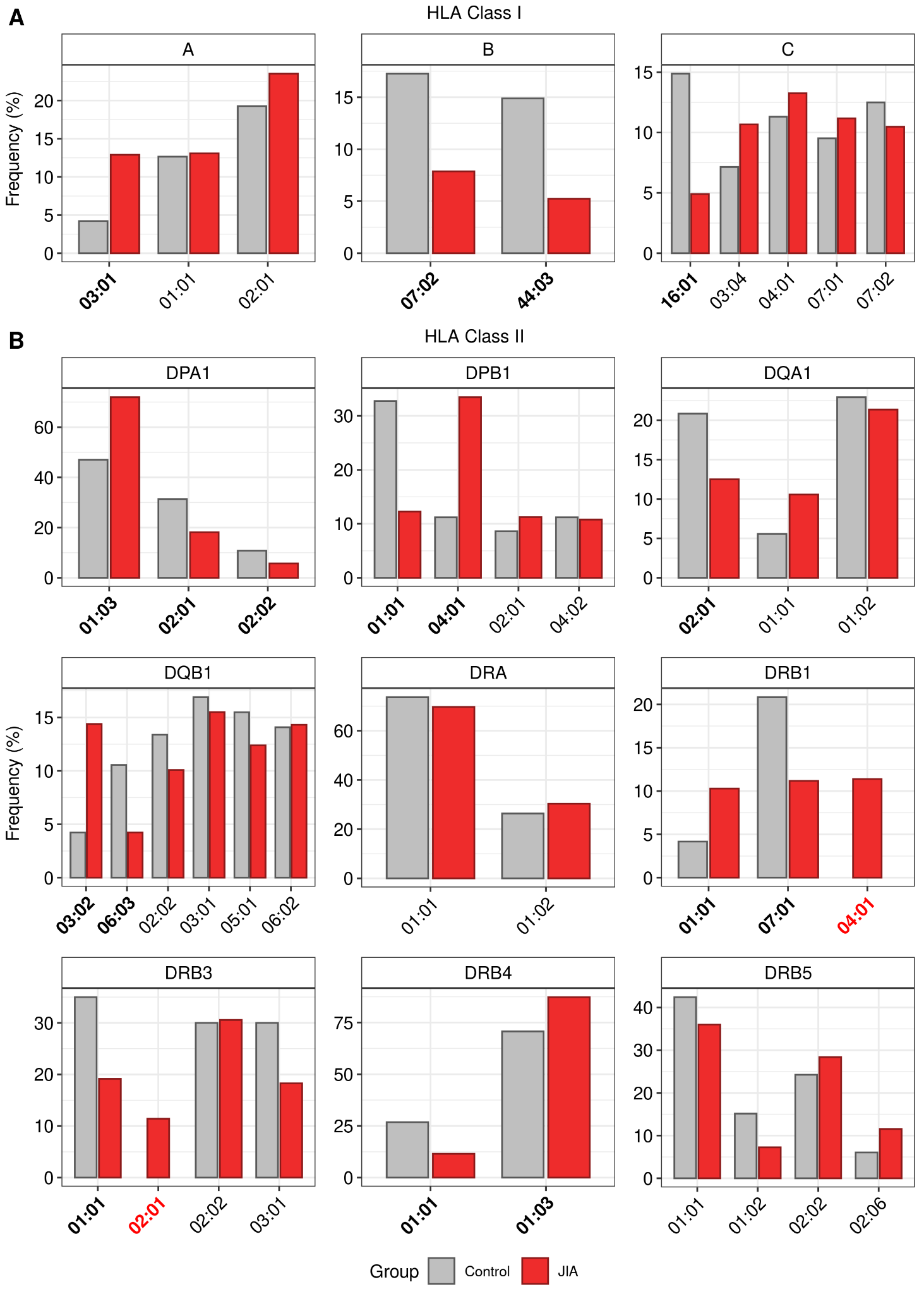

**Supplementary Figure S8. Consensus frequencies of HLA allele types in healthy control subjects and JIA patients.** (**A**) Bar plots showing the consensus frequencies of HLA allele types at HLA class I loci. (**B**) Bar plots showing the consensus frequencies of HLA allele types at HLA class II loci. The x- and y-axis indicate the names and frequencies of HLA allele types, respectively. The names of HLA types detected only in the JIA group are marked in red. The names of significantly different HLA types observed in the healthy control and JIA groups are represented in bold (P < 0.05).

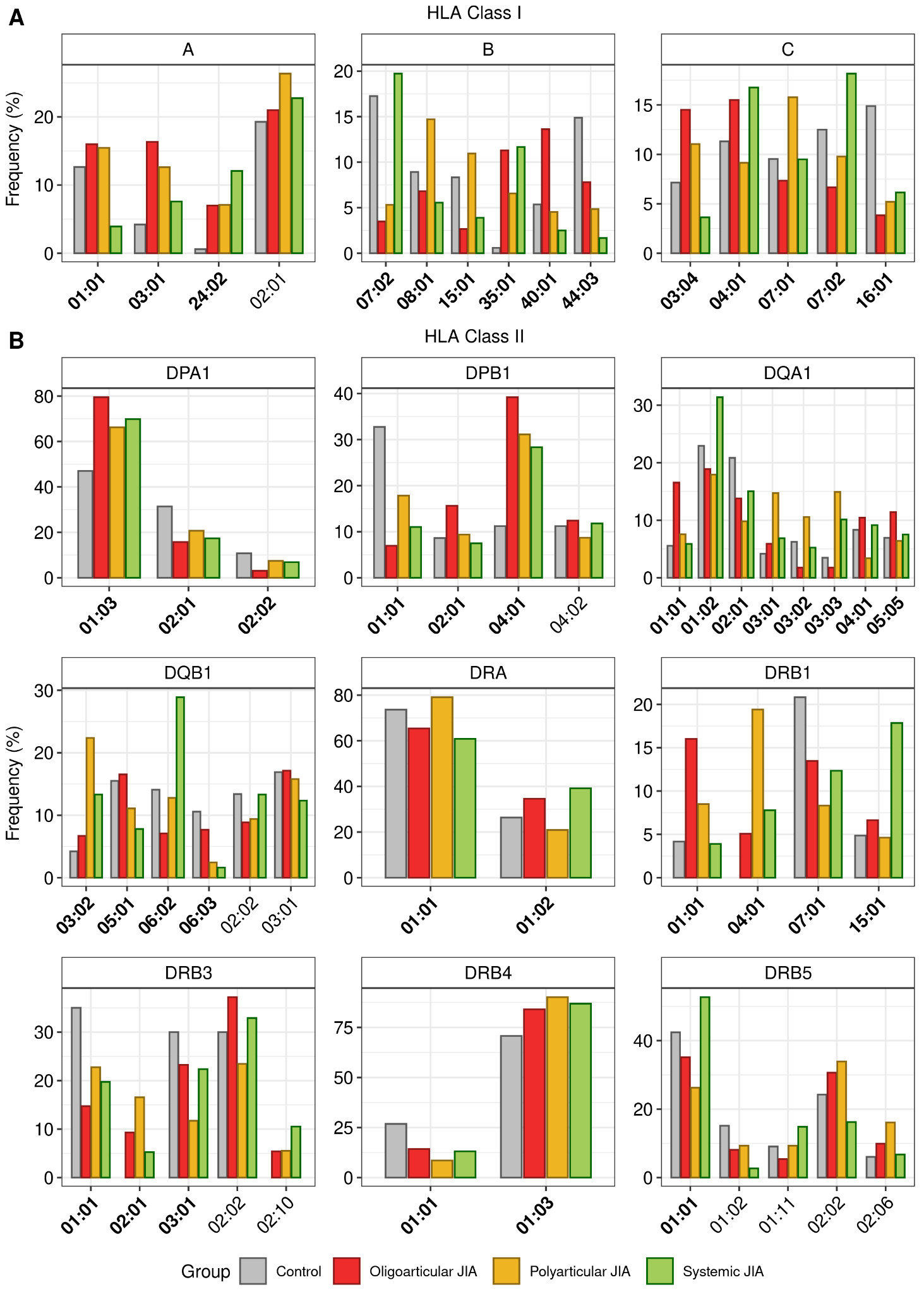

**Supplementary Figure S9. Consensus frequencies of HLA allele types in healthy control subjects and JIA patients grouped by subtypes.** (**A**) Bar plots showing the consensus frequencies of HLA allele types at HLA class I loci. (**B**) Bar plots showing the consensus frequencies of HLA allele types at HLA class II loci. The x- and y-axis indicate the names and frequencies of HLA allele types, respectively. The names of significantly different HLA types observed in the healthy control and 3 JIA subtype groups are represented in bold (P < 0.05).

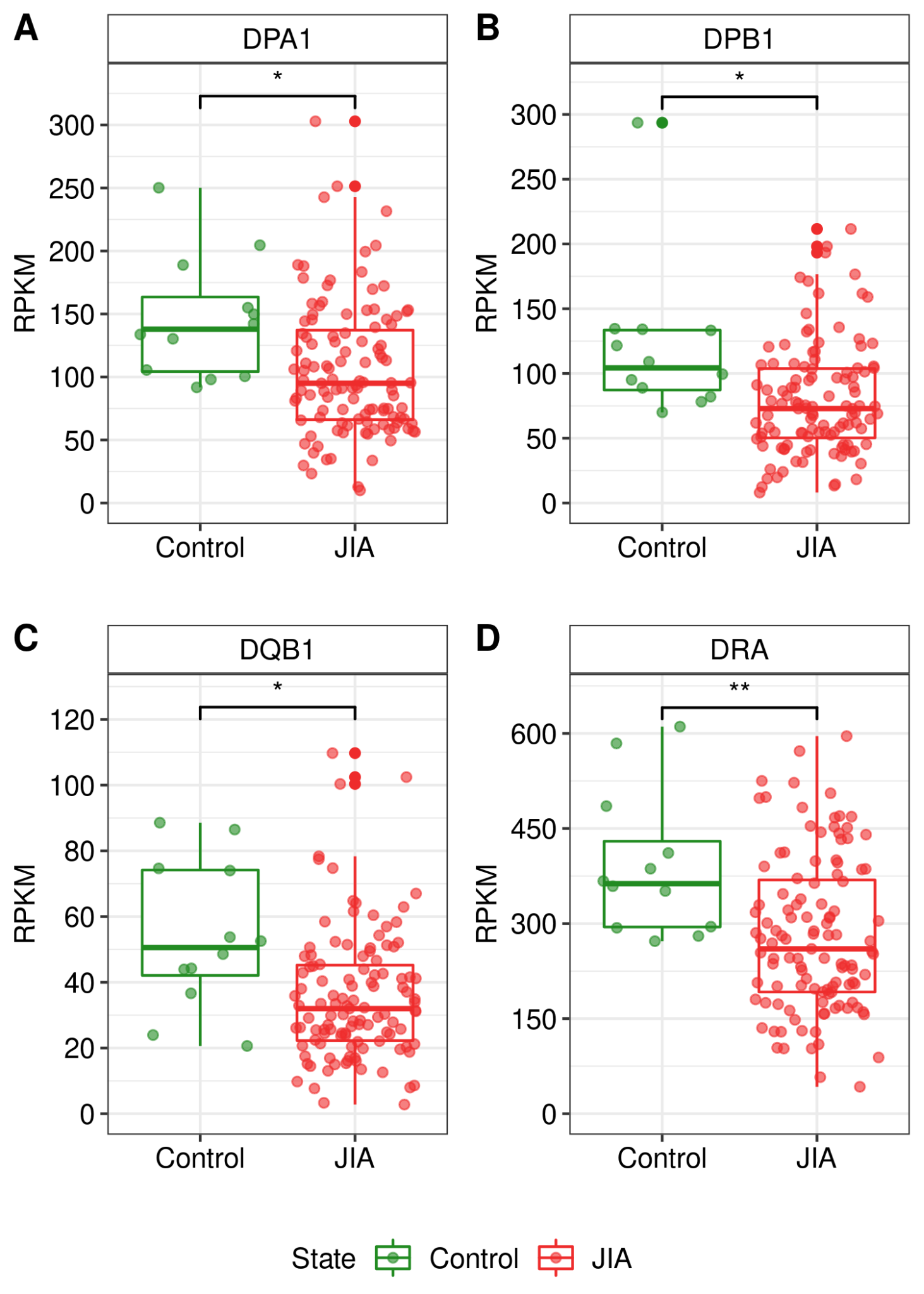

**Supplementary Figure S10. Boxplots showing the locus-specific expression levels of HLA class Ⅱ genes.** Boxplots showing the expression levels of HLA class Ⅱ genes, (**A**) *DPA1*, (**B**) *DPB1*, (**C**) *DQB1*, and (**D**) *DRA*, in healthy controls and JIA patients. Asterisks represent the significance levels of difference between the HLA gene expression levels in JIA patients and those in healthy controls. *, P < 0.05; **, P < 0.01.

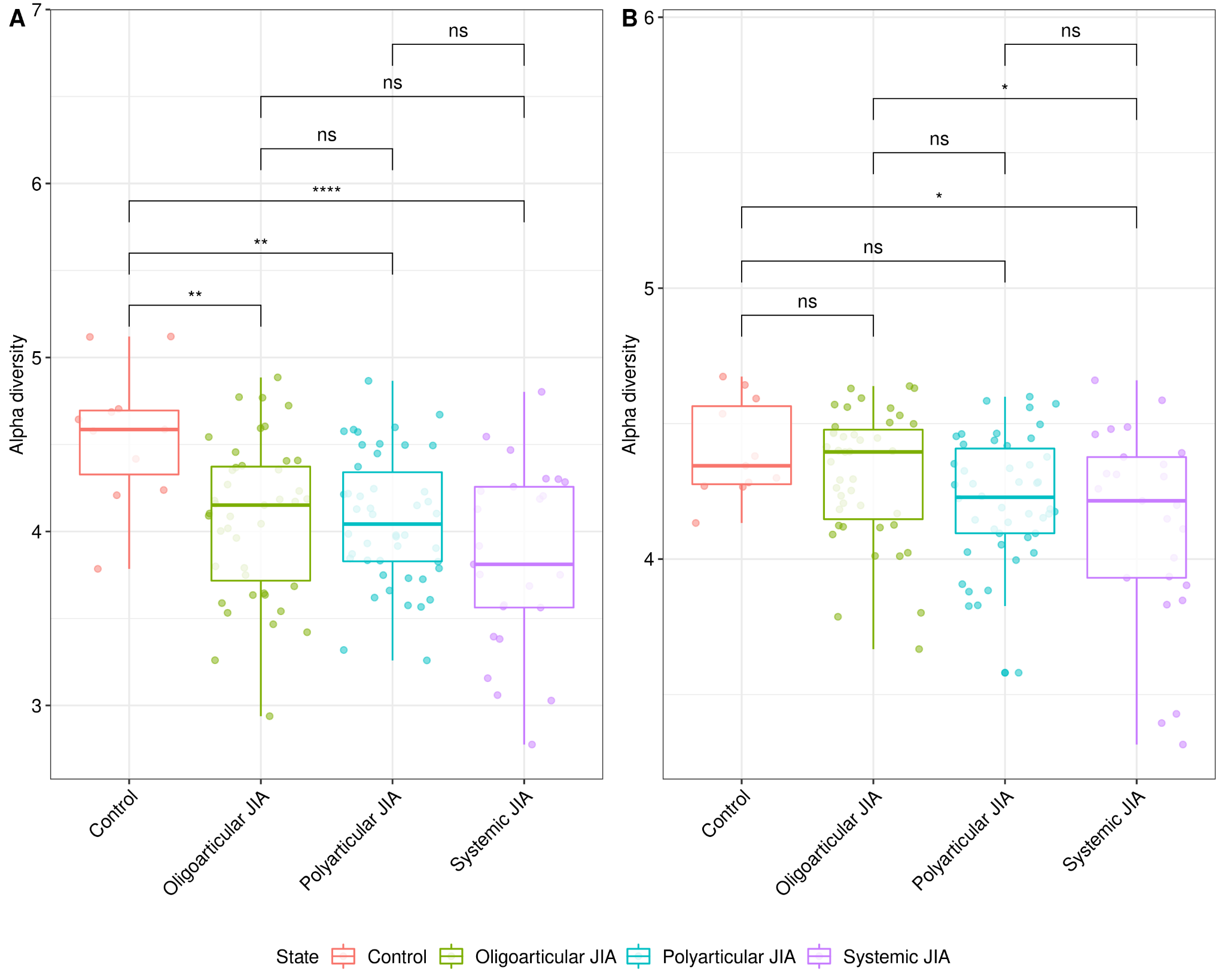

**Supplementary Figure S11. The clonotypic diversities of TCRA and TCRB loci in healthy controls and patients of 3 JIA subtypes.** Box plots showing the alpha diversities of (**A**) TCRA and (**B**) TCRB loci in the healthy controls and 3 JIA subtypes. The y-axis indicates alpha diversity representing the clonotypic diversity. Colored dots indicate samples of healthy controls and JIA patients of specific subtypes, respectively. Asterisks represent the significance levels of differences between the clonotypic diversities of TCRA and TCRB in healthy controls and those in patients of each JIA subtype.

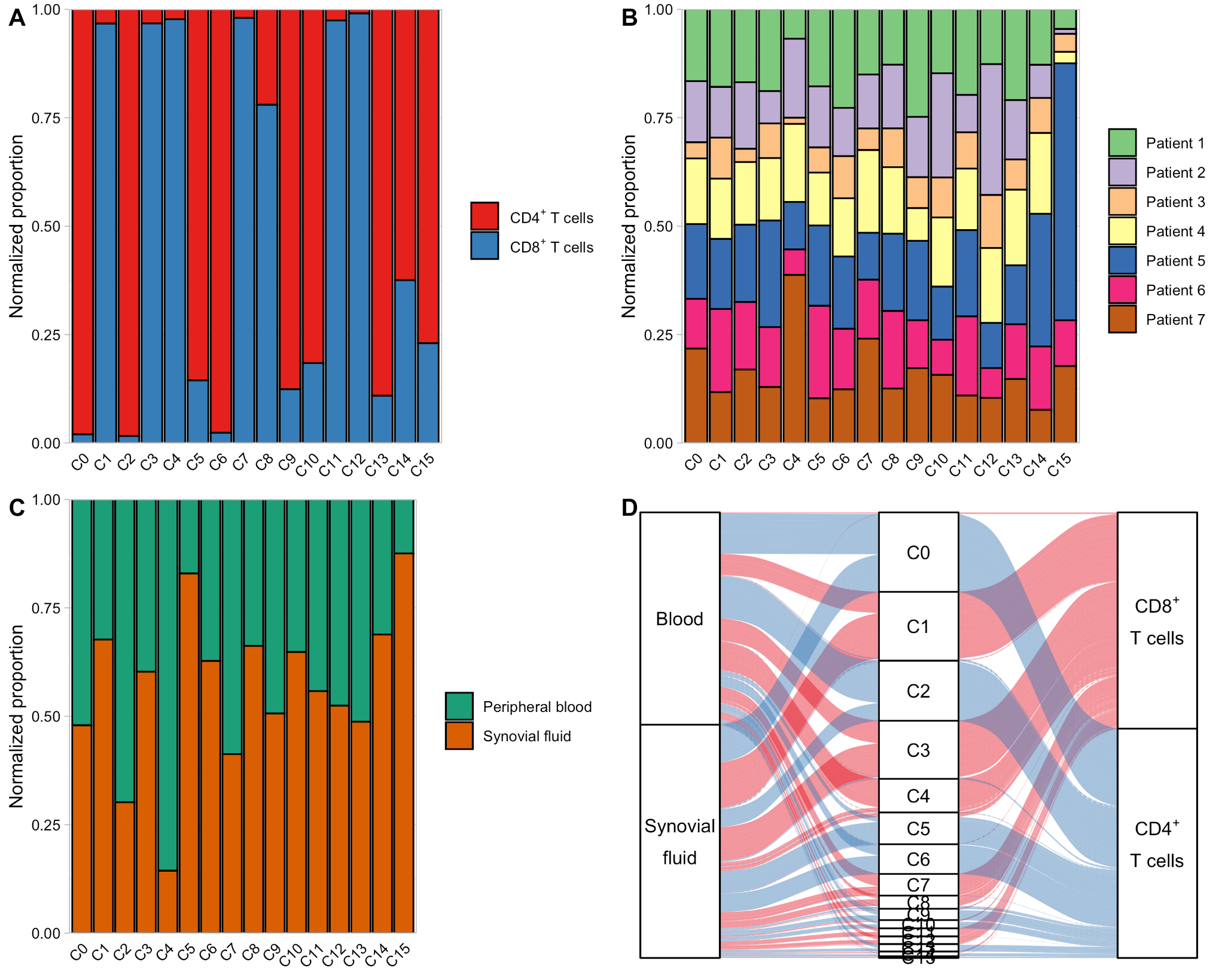

**Supplementary Figure S12. Quantification of the cluster distribution.** (**A**) Bar plot showing cluster distribution of the two different T cell types. (**B**) Bar plot showing cluster distribution of the seven different JIA patients. (**C**) Bar plot showing cluster distribution of the two tissue types. The y-axis of the bar graph denotes the normalized proportions. (**D**) Alluvial diagram showing the distribution of T cell types. Three categorical axes are variables (i.e., origin tissues, clusters, and T cell types) along which the data are grouped. Each horizontal spline (called alluvium) corresponds to a fixed value of each axis variable, indicated by T cell types’ of colors. The red and blue colors correspond to CD8^+^ and CD4^+^ T cells, respectively.

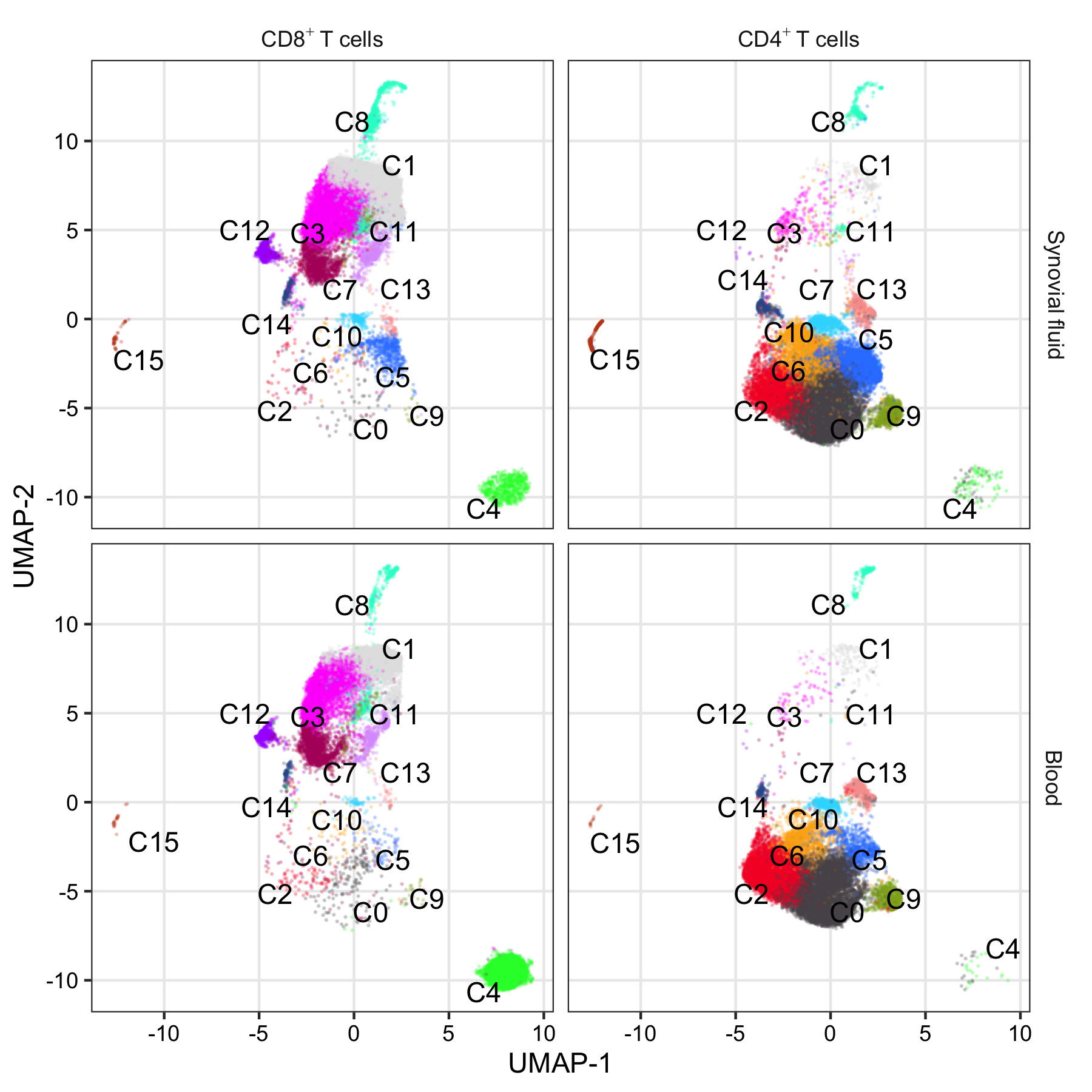

**Supplementary Figure S13. UMAP plot for T cell types and tissue types in different clusters.** Each dot represents an individual T cell and color indicates cluster origin.

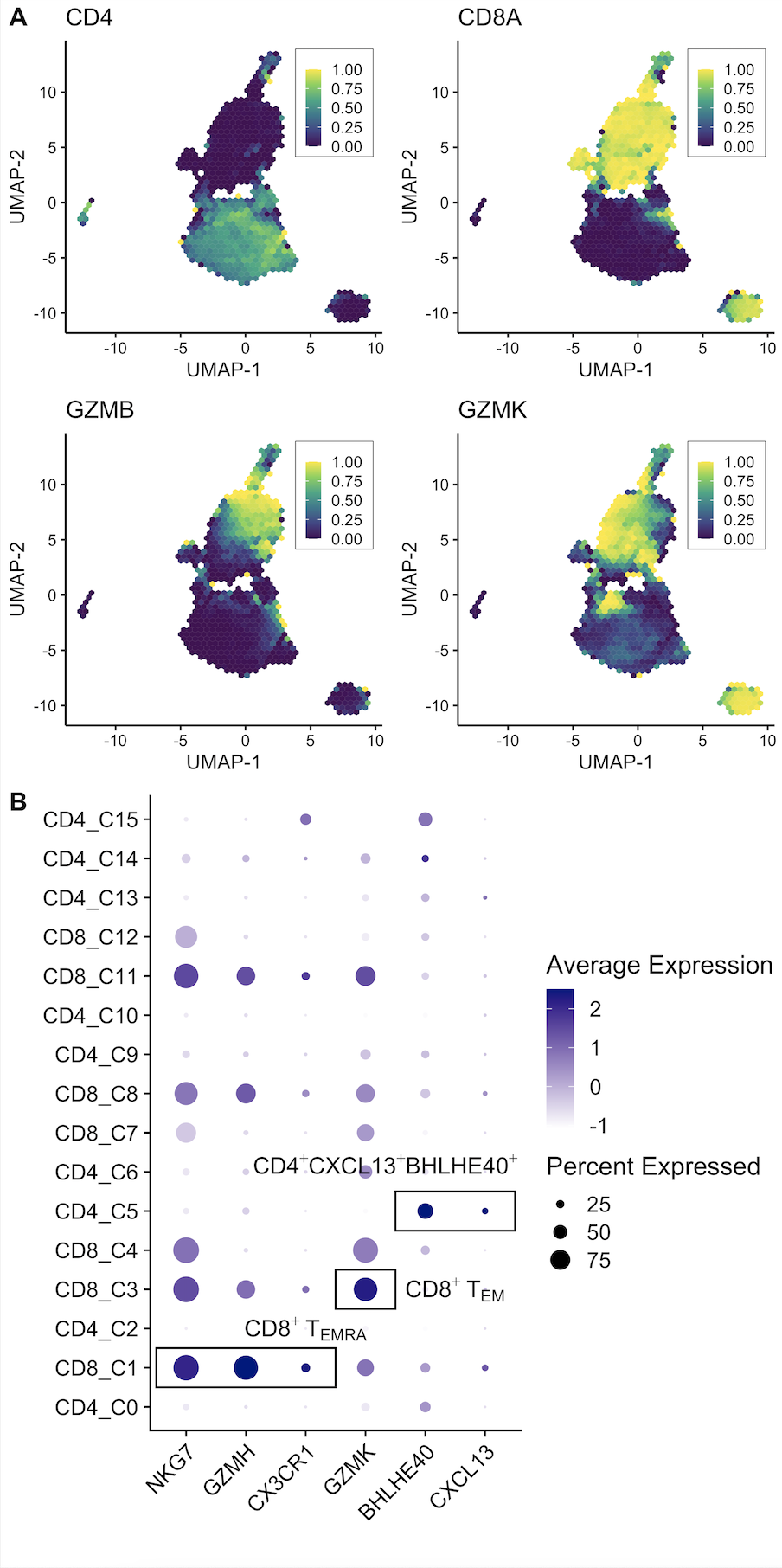

**Supplementary Figure S14. Expression levels of signature genes in each cluster.** (**A**) UMAP plot showing the expression levels of four selected genes. Each hexagon represents summarizing points into binned hexagon cells (The number of bins partitioning the range = 50) using schex R package. The color bar denotes the proportion of observations in the bin greater than 0. (**B**) Dot plot representing the expression levels of six selected genes. The size of dots denotes the percentage of cells within a cluster and the color bar encodes the average expression level of the selected genes across all cells within a cluster.

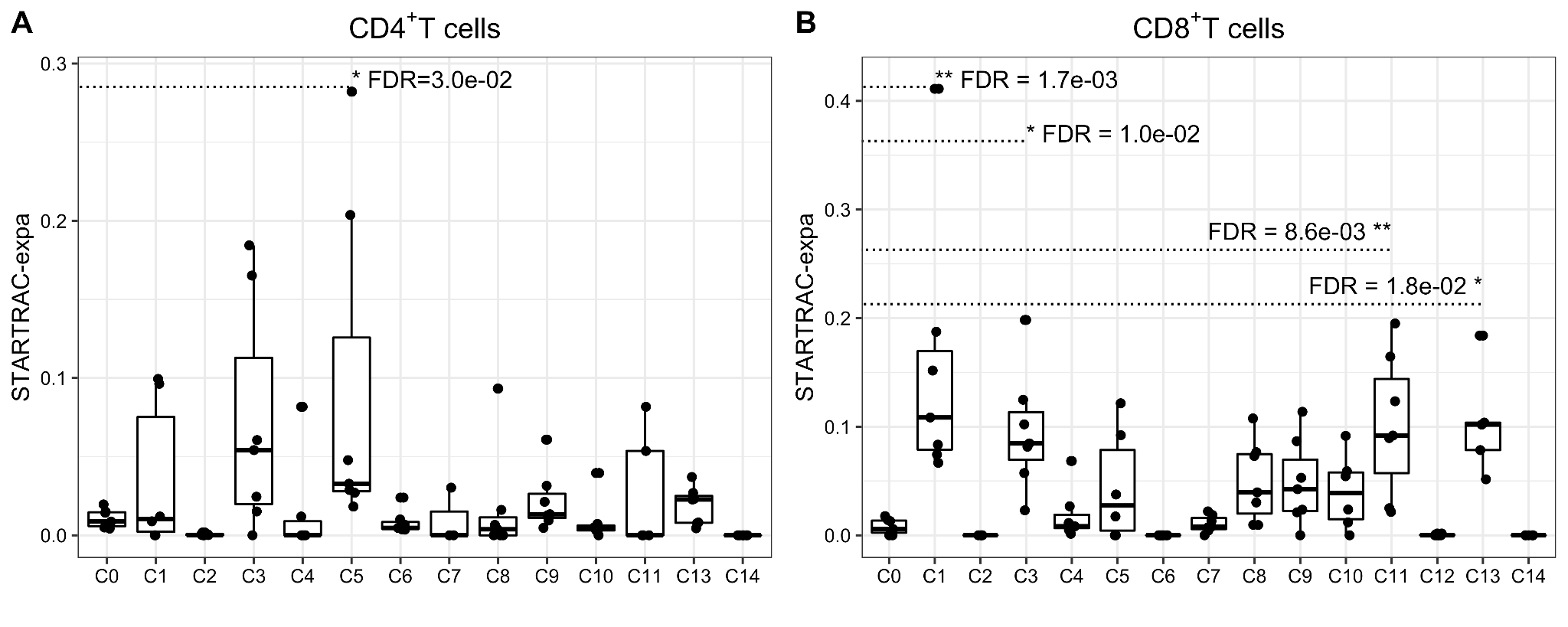

**Supplementary Figure S15. Clonal expansion levels of the clusters.** Clonal expansion levels of each cluster in (**A**) CD4^+^ T cells and (**B**) CD8^+^ T cells. STARTRAC-expa quantified clonal expansion levels for each JIA patient (n = 7). *FDR < 0.05, **FDR < 0.01, two-sided Wilcoxon test.

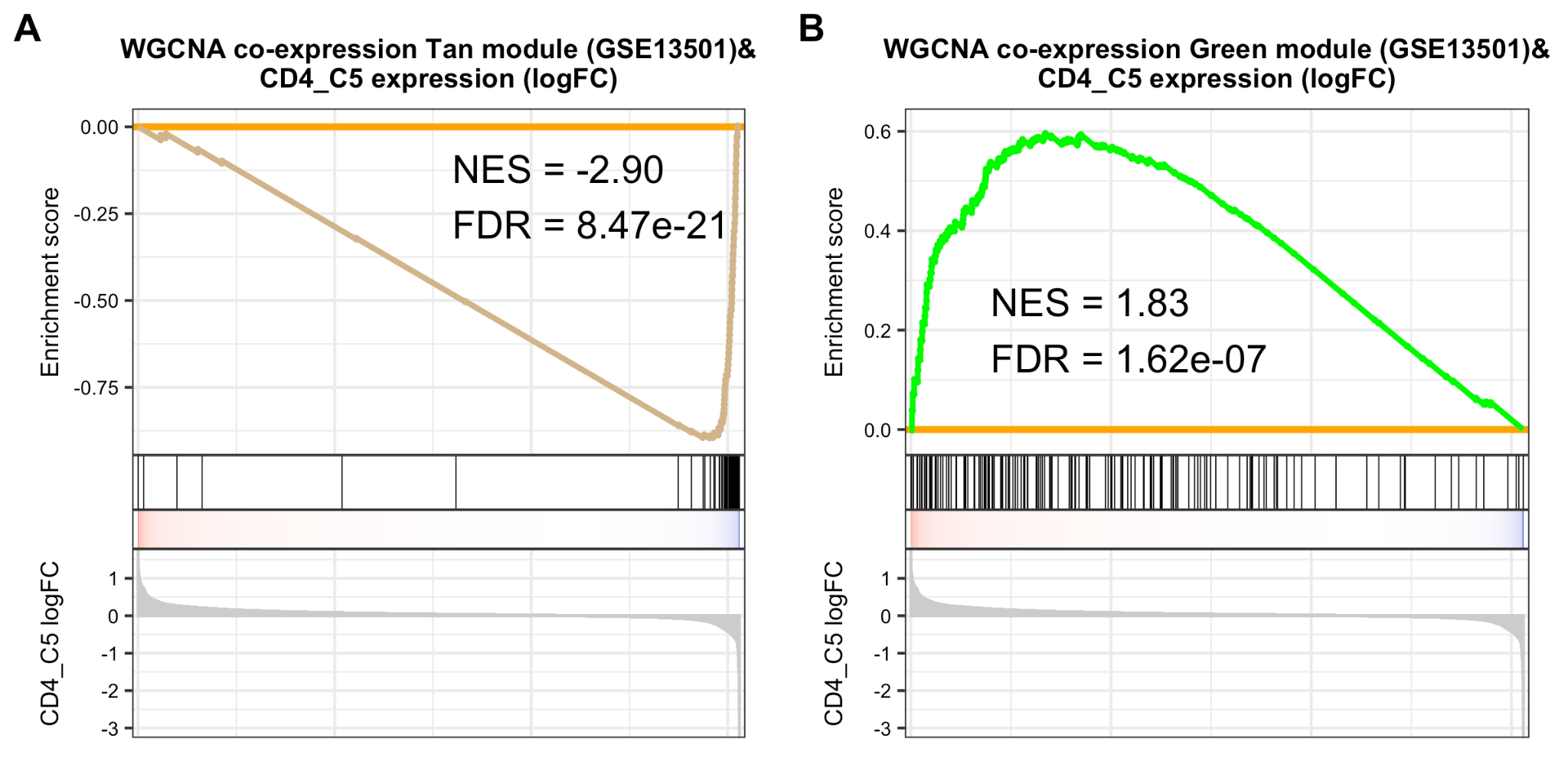

**Supplementary Figure S16. Module enrichment analysis between expression levels of scRNA-seq CD4_C5 cluster genes and co-expression gene sets derived from JIA case-control expression data.** GSEA plots between expression levels of scRNA-seq CD4_C5 cluster and co-expression of (**A**) Tan and (**B**) Green modules derived from case-control expression data of GSE13501. The gene lists of modules are in Table S7.
